# Assessment of Nirmatrelvir-Ritonavir Effects on Acute and Post-Acute COVID-19 Illness in US Adolescents: Target Trial Emulation

**DOI:** 10.1101/2025.04.02.25324861

**Authors:** Yishan Shen, Bingyu Zhang, Ravi Jhaveri, Jiayi Tong, Ting Zhou, Jiajie Chen, Dazheng Zhang, Qiong Wu, Chengxi Zang, Fei Wang, Rainu Kaushal, Jason G. Newland, Emily Taylor, Sharon J. Herring, Yuriy Bisyuk, David Liebovitz, Susan Kim, Daniel Fort, David A. Williams, Benjamin D. Horne, Mollie R. Cummins, Mei Liu, Elizabeth A. Chrischilles, Lindsay G. Cowell, Abu Saleh Mohammad Mosa, Payal Patel, Christopher B. Forrest, Yong Chen, the RECOVER Consortium

## Abstract

**IMPORTANCE:** Nirmatrelvir-ritonavir, an efficacious treatment for acute COVID-19, has yielded conflicting evidence regarding its effectiveness in preventing Long COVID among adults. The effectiveness of nirmatrelvir-ritonavir in adolescents in real-world settings is not well understood.

**OBJECTIVE:** To evaluate the effectiveness of nirmatrelvir-ritonavir in adolescents aged 12 to 20 years given within the first five days of acute COVID-19 illness on illness severity during the acute (days 0 to 27) and post-acute phases (days 28 to 179).

**DESIGN, SETTING, AND PARTICIPANTS:** This target trial emulation used the electronic health records (EHR) data from twenty-nine medical institutions participating in the NIH’s RECOVER consortium. We included data collected between April 1, 2022 and December 31, 2023, and included all patients with a SARS-CoV-2 infection who were not hospitalized on the day of entry. Those who were treated with oral nirmatrelvir-ritonavir within 5 days after the positive test (n=2,923) and those who did not received nirmatrelvir-ritonavir within 5 days after the positive test during the acute phase of SARS-CoV-2 infection (control group, n=31,947) were identified.

**EXPOSURES:** Treatment with nirmatrelvir-ritonavir based on prescription records.

**MAIN OUTCOMES AND MEASURES:** We used modified Poisson regression models for binary outcomes to examine the effects of nirmatrelvir-ritonavir on acute phase hospitalization, emergency department (ED) visits, and outpatient visits as well as progression to moderate or severe acute illness among individuals treated with nirmatrelvir-ritonavir compared to control counterparts. Additionally, we assessed the medication’s associations with 16 conditions and symptoms during the acute phase of illness and its subsequent association with specific Long COVID diagnostic code U09.9 during the post-acute phase. We reported the absolute risks and estimated the relative risks (RRs), with adjustments made for demographic variables, clinical characteristics, and healthcare utilization, using propensity score matching.

**RESULTS:** The study cohort included 34,870 adolescents (median [Q1, Q3] age 16.0 [14.0, 18.0]); 46.0% male and 43.2% Non-Hispanic White. Among these patients, 2,923 were treated with nirmatrelvir-ritonavir within 5 days of cohort entry and 31,947 received no treatment. The absolute rates were 0.58% vs. 0.96% for hospitalization, 74.17% vs. 82.66% for outpatient visits, 1.81% vs. 2.28% for ED visits, 3.08% vs. 3.99% for moderate/severe acute illness, and 0.21% vs. 0.20% for long COVID diagnosis in the treatment vs. control group. After propensity score matching, during acute-phase period, compared to the control group, nirmatrelvir-ritonavir was associated with reduced risk of any hospitalization (relative risk (RR) 0.48; 95% confidence interval (CI) 0.29-0.80), outpatient visit (RR 0.86; 95% CI 0.82-0.90), and moderate/severe acute illness (RR 0.69; 95% CI 0.56-0.87). Additionally, nirmatrelvir-ritonavir was associated with reduced risk of 7 of 16 specified conditions in chest pain (RR 0.41; 95% CI 0.26-0.63), fatigue and malaise (RR 0.49; 95% CI 0.32-0.76), generalized pain (RR 0.65; 95% CI 0.45-0.95), headache (RR 0.60; 95% CI 0.44-0.83), mental health (RR 0.56; 95% CI 0.39-0.80), musculoskeletal (RR 0.63, 95% CI 0.43-0.94), and respiratory signs and symptoms (RR 0.61, 95% CI 0.52-0.71). No evidence was found to suggest that the treatment was associated with the risk of long COVID diagnosis U09.9 (RR 0.96, 95% CI 0.40-2.32).

**CONCLUSIONS AND RELEVANCE:** This study found that treatment with nirmatrelvir-ritonavir in adolescents within 5 days of SARS-CoV-2 infection, was associated with reduced risk of acute-phase COVID-19 illness and subsequent healthcare utilization. However, treatment was not associated with a lower risk of Long COVID diagnosis.

**Key Points:** *Question:* Does treatment with nirmatrelvir-ritonavir reduce the acute COVID-19 illness severity and the risk of developing Long COVID in adolescents aged 12 to 20 years?

*Findings:* In this target trial emulation study of 34,870 adolescents with SARS-CoV-2 infection who were not hospitalized on the cohort entry day, 2,923 who were treated with nirmatrelvir-ritonavir exhibited lower acute illness severity including reduced risk of 7 of 16 specified conditions and symptoms, as well as reduced risk of acute-phase hospitalization and outpatient visits, compared with 31,947 who received no treatment. The association with reduced risk of Long COVID was not observed in this study.

*Meaning:* In adolescents, nirmatrelvir-ritonavir appears to be effective at reducing the severity of acute COVID illness but was not associated with the risk of developing Long COVID.

## Introduction

The COVID-19 pandemic has prompted rapid development and evaluation of antiviral therapies to mitigate disease severity and progression. Nirmatrelvir-ritonavir, an oral antiviral combination, has emerged as an efficacious treatment for SARS-CoV-2 infection in adults according to clinical trials^1^, and yielded mixed evidence regarding its effectiveness in preventing Long COVID. However, limited data exist on its effects in adolescents, particularly regarding acute and post-acute sequelae of SARS-CoV-2 (PASC, or “long COVID”).

In adult populations, nirmatrelvir-ritonavir has shown significant benefits for acute COVID-19 illness. Randomized clinical trials by Hammond et al reported 89% reduction in the risk of COVID-19–related hospitalization or death among high-risk, unvaccinated adults with COVID-19^1^, although it was not associated with a significantly shorter time to sustained alleviation of COVID-19 symptoms in vaccinated or unvaccinated outpatients^2^. Real-world evidence also suggests that nirmatrelvir-ritonavir reduces the risk of progression to severe disease, hospitalization, and death, particularly in unvaccinated or high-risk adult patients^3–5^. However, there is mixed evidence regarding nirmatrelvir-ritonavir’s potential impact on reducing long COVID risk. While some reports suggest a potential association between nirmatrelvir-ritonavir use and decreased long COVID risk^6–11^, conflicting results^6,12^ in the same population have arisen from studies utilizing the healthcare databases of the US Department of Veterans Affairs (VA). Some studies found no association between nirmatrelvir-ritonavir use and the risk of long COVID^13–15^, and a recent randomized clinical trial reported no significant benefit in improving selected long COVID symptoms^16^.

Research on nirmatrelvir-ritonavir’s effectiveness in younger populations, particularly adolescents, remains limited, with most clinical effectiveness data comes from adult populations. While the drug is recommended for adolescents with risk factors for severe disease^17–19^, pharmacokinetic and pharmacodynamic data are lacking, and evidence on its use, safety, and effectiveness in this population is scarce^20^. Adolescents may exhibit different immune responses and clinical outcomes compared with adults^21,22^, raising questions about the generalizability of adult trial results although a phase 2/3 clinical trial assessing the safety and efficacy of nirmatrelvir-ritonavir treatment in children 6 to 17 years of age with COVID-19 is currently underway^23^. The potential role of nirmatrelvir-ritonavir in preventing acute COVID-19 severity and long COVID, which may manifest differently in adolescents than in adults, remains largely underexplored. Recent studies have reported associations between nirmatrelvir-ritonavir treatment and specific long COVID conditions among adolescents^24^. Additionally, a recent target trial emulation found that this treatment was associated with reduced 28-day all-cause hospitalization among non-hospitalized pediatric patients aged 12-17 years with SARS-CoV-2 Omicron variant infection^25^. However, these did not evaluate the potential impact of nirmatrelvir-ritonavir on long COVID within this population. These findings underscore the need for further more comprehensive research to elucidate whether this therapy mitigates acute COVID-19 severity and reduces the risk of developing long COVID among pediatric populations.

In this study, we emulated a target trial to investigate the effectiveness of nirmatrelvir-ritonavir on acute and post-acute COVID-19 outcomes among adolescents aged 12 to 20 years in the United States. Through the study leveraging data from the RECOVER consortium, which consists of twenty-nine US children’s hospitals and health institutions, we aim to provide crucial insights into the effectiveness of nirmatrelvir-ritonavir in reducing acute COVID-19 severity, preventing hospitalizations, emergency department visits, and outpatient visits, as well as mitigating long-term complications in adolescents. This study boasts several strengths. First, it is the first and largest study to date that evaluates the effectiveness of nirmatrelvir-ritonavir across both the acute and post-acute phases in adolescents. Second, it encompasses a comprehensive assessment of a wide array of outcomes. Additionally, this study extends the follow-up period beyond that of previous pediatric studies, providing a more longitudinal perspective on the impacts of the treatment. Finally, rigorous sensitivity analyses and negative control experiments were conducted to ensure the robustness and reliability of our clinical findings. Collectively, these strengths underscore the significance of our research, which aims to guide clinical decision-making and enhance public health strategies for managing COVID-19 among adolescents.

## Methods

### Data Sources

This study is part of the National Institutes of Health (NIH) funded RECOVER Initiative (https://recovercovid.org/), which aims to learn about the long-term effects of COVID-19. This study included twenty-nine US children’s hospitals and health institutions. The EHR data were standardized to the PCORnet Common Data Model (CDM) and extracted from the RECOVER Database Version s11. More details are available in the Supplementary Materials (Section S1).

### Cohort Construction

We conducted a target trial emulation with patient entry date from April 1, 2022, to December 31, 2023. SARS-CoV-2 infections were defined by positive PCR and antigen tests, diagnoses of COVID-19, or a prescription to be treated with nirmatrelvir-ritonavir (in cases lacking a viral test or diagnosis code record). The index date (i.e., cohort entry date) was set as either the earliest date of a positive viral test, COVID-19 diagnosis, or nirmatrelvir-ritonavir was prescribed in instances where the first two were not documented. If any prescription of nirmatrelvir-ritonavir was noted 3 days prior to the recorded date of a positive viral test or COVID diagnosis, the index date was adjusted to the date the medication was prescribed.

We included adolescents who aged between 12 and 20 years old^26^, weighed over 40 kg, who were in an outpatient setting, who were not hospitalized on the day of cohort entry, and who had at least one healthcare visit within the baseline period (24 months to 7 days before the index date). Patients diagnosed with multisystem inflammatory syndrome in children (MIS-C), chronic kidney disease, end-stage kidney disease, hepatic failure or insufficiency, or procedure with chronic dialysis or kidney transplant, or had contraindication medications up to 90 days before cohort entry^27^, were excluded from this study.

The treated group included participants with SARS-CoV-2 infection with an index date from April 1, 2022, to December 31, 2023, and prescribed nirmatrelvir-ritonavir within 5 days of the index date. The control group was constructed from the patients who had SARS-CoV-2 infection from April 1, 2022, to December 31, 2023, and were not prescribed nirmatrelvir-ritonavir within 5 days of the index date, regardless of if they used any other outpatient COVID-19 antiviral or antibody treatments.

### Outcomes

The outcomes of interest in our study include the clinical indicators of COVID-19 acute illness severity and healthcare utilization (hospitalizations, outpatient visits, and emergency department (ED) visits) during the acute phase (0-27 days following cohort entry).

Additionally, our study includes a comprehensive assessment of 16 various conditions and symptoms within the acute phase reviewed by pediatric physicians^28^, such as abdominal pain, arrhythmias, cardiovascular signs and symptoms, chest pain, fatigue and malaise, fever and chills, fluid and electrolyte imbalances, generalized pain, headache, mental health issues, musculoskeletal issues, respiratory signs and symptoms, skin conditions, diarrhea, nausea, and vomiting.

We also investigated the impact of nirmatrelvir-ritonavir during the post-acute phase (28-179 days following cohort entry). We used ICD-10-CM code, U09.9, specific for post-COVID-19 conditions to indicate long COVID. For this, we created a binary outcome, which was defined as positive if anyone was identified as having long COVID with a U09.9 diagnosis code.

These outcomes were assessed during the follow-up period in patients without a history of the specific condition during the baseline period. We used validated diagnostic codes (ICD10CM, ICD10, ICD9CM, SNOMED) confirmed by two board-certified pediatricians (RJ, CF), with details of the code sets available in the **Supplementary Table S2**.

### Covariates

To account for potential confounding bias^29^, an extensive set of confounding variables^30^ was incorporated including demographic factors, including patient age at index date (in year), sex (female, male), and race and ethnicity (Non-Hispanic White (NHW), Non-Hispanic Black (NHB), Hispanic, Asian American/Pacific Islander (AAPI), Multiple, Other/Unknown); clinical factors, including obesity status, a chronic condition indicator as defined by the Pediatric Medical Complexity Algorithm^31^ no chronic condition, non-complex chronic condition, complex chronic condition), and a list of pre-existing chronic conditions^32^; healthcare utilization factors collected 24 months to 7 days before index date, including the number of inpatient visits, outpatient visits, ED visits, unique medications, and negative COVID-19 tests (0, 1, 2, ≥3); vaccine information, including the number of COVID-19 vaccine doses before index date (0, 1, ≥2) and interval since the last COVID-19 immunization date (no vaccine, <4 months, ≥ 4 months); year-month of cohort entry (from April 2022 to December 2023); indicators from the data-contributing sites. The detailed definitions of study variables were included in **Supplementary Table S2**.

### Statistical Analysis

To mitigate the potential effects of confounding variables, we employed a propensity score model to adjust for a wide array of measured confounders identified prior to the index date. This step involved fitting a logistic regression model, where the treatment status was regressed on the study variables listed as covariates. The predicted probabilities obtained via the logistic regression model yield the propensity score for each patient. Following the 1:5 ratio matching^33,34^ of treated to untreated subjects, we evaluated the standardized mean difference (SMD) for each covariate between the treated vs not treated groups. An SMD of 0.1 or less was considered indicative of a satisfactory balance^35^.

We used a modified Poisson regression model for binary outcome^36^ to calculate the relative risk (RR) between the two comparison groups for each outcome, as we did in our earlier investigation^30,37–39^.

To evaluate the robustness of findings across different subgroups, we also conducted subgroup analyses by age (12-15, 16-20 years), obesity status (yes, no), vaccination status (yes, no), health risk groups (no, at least one risk factor), and the severity of COVID-19 during the acute phase. The risk factors included obesity, cardiovascular disease, asthma, hypertension, diabetes, neurological disease, genetic/metabolic disease, and cancer. Acute COVID-19 severity^40^ was categorized as asymptomatic, mild (symptomatic), moderate (involving moderately severe COVID-19-related conditions like gastroenteritis, dehydration, and pneumonia), and severe (comprising unstable COVID-19-related conditions, ICU admission, or mechanical ventilation). These were then grouped into two categories: “non-severe” (asymptomatic) and “severe” (mild, moderate, and severe).

### Sensitivity Analysis

Extensive sensitivity analyses were conducted to evaluate the robustness of the research findings; see Sections S4-6 of the Supplementary Materials. In the primary analysis, we focused on adolescents aged 12 to 20. To further examine the effectiveness of nirmatrelvir-ritonavir in both children and adolescents, we included a sensitivity analysis that included patients aged under 12, while keeping all other inclusion and exclusion criteria the same (Section S4). Another sensitivity analysis was conducted to relax the weight criteria, i.e., including adolescents whose weights were under 40 kg or missing (Section S5).

In target trial emulation studies, residual study bias could exist from unmeasured and systematic sources even after controlling for measured confounders. In response, we conducted a sensitivity analysis of negative control outcome (NCO) experiments^41–43^. In this NCO analysis, the null hypothesis of no effect was believed to be true using 36 negative control outcomes prespecified by pediatric physicians (Section S6). The empirical null distribution and calibrated effectiveness were reported and detailed in Section S6 of the Supplementary Materials.

## Results

### Cohort Identification

Within the RECOVER network, we identified a total of 34,870 participants, 2,923 treated and 31,947 untreated with nirmatrelvir-ritonavir, who ranged in age from 12 to 20 years, encompassing a diverse representation of sex and race/ethnicity. **Figure 1** shows a flowchart of the construction of the cohort as detailed in Methods. Among these participants, males constituted 46.0% of the cohort, while females represented 54.0%. The distribution across race/ethnic groups was varied with 24.6% Hispanic, 43.2% NHW, 15.7% NHB, 5.2% AAPI, and 1.3% identifying as Multiple. The baseline characteristics stratified by the treatment group and the control group are detailed in **Table 1**.

**Figure 1.**
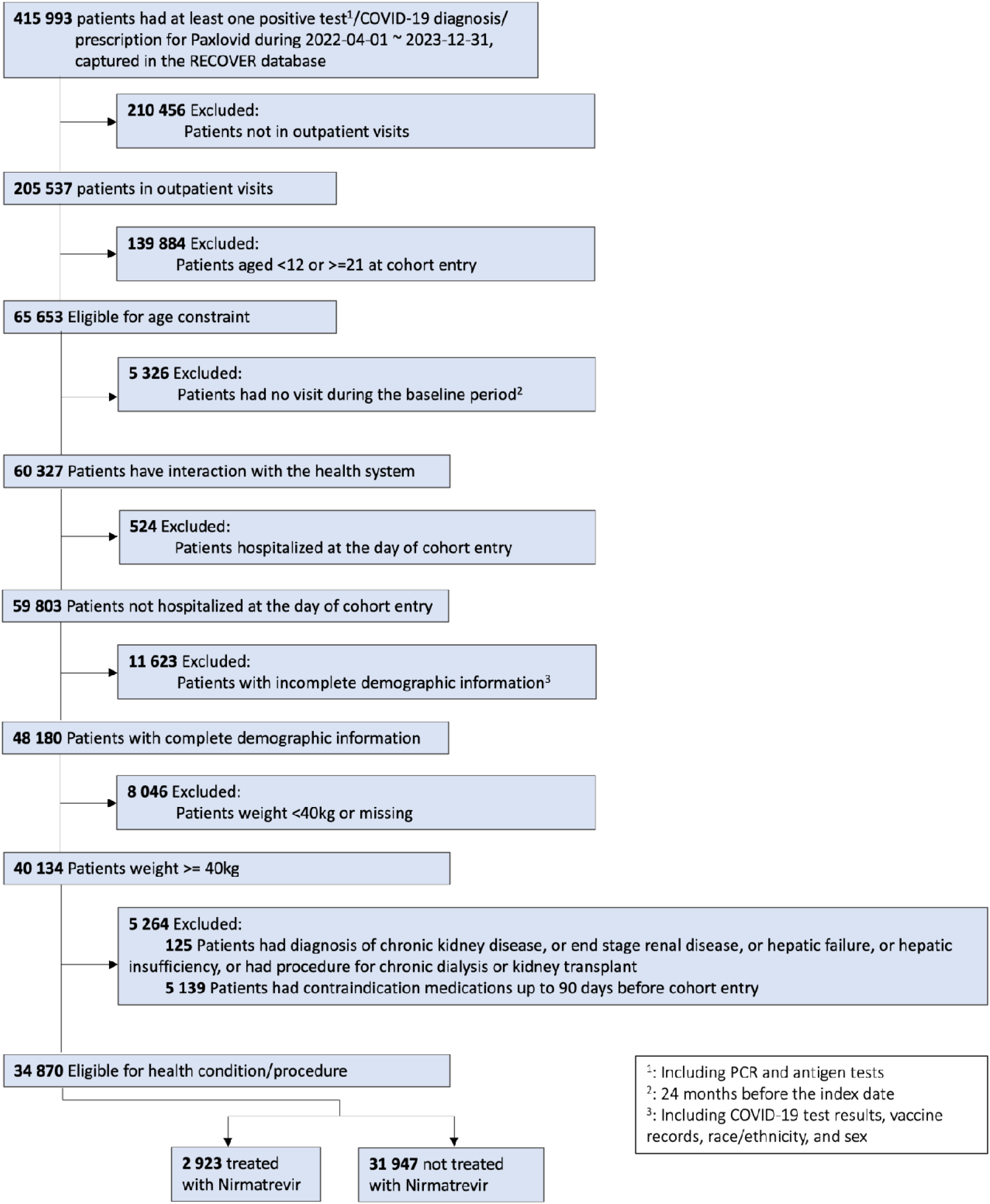
Flowchart of construction of the treatment and control groups.

**Table 1.** Baseline characteristics of participants of nirmatrelvir-ritonavir versus no treatment.

|  | Treated with<br>nirmatrelvir-ritonavir<br>(N=2,923) | Not treated<br>with<br>nirmatrelvir-ritonavir<br>(N=31,947) | Overall<br>(N=34,870) |
| --- | --- | --- | --- |
| **Median Age [Q1, Q3]** |  |  |  |
|  | 17.0 [15.0, 19.0] | 16.0 [14.0, 18.0] | 16.0 [14.0, 18.0] |
| **Age** |  |  |  |
| 12 | 142 (4.9%) | 2885 (9.0%) | 3027 (8.7%) |
| 13 | 199 (6.8%) | 3472 (10.9%) | 3671 (10.5%) |
| 14 | 253 (8.7%) | 3781 (11.8%) | 4034 (11.6%) |
| 15 | 289 (9.9%) | 4128 (12.9%) | 4417 (12.7%) |
| 16 | 383 (13.1%) | 4072 (12.7%) | 4455 (12.8%) |
| 17 | 392 (13.4%) | 3943 (12.3%) | 4335 (12.4%) |
| 18 | 423 (14.5%) | 3119 (9.8%) | 3542 (10.2%) |
| 19 | 412 (14.1%) | 3179 (10.0%) | 3591 (10.3%) |
| 20 | 430 (14.7%) | 3368 (10.5%) | 3798 (10.9%) |
| **Sex** |  |  |  |
| **Female** | 1530 (52.3%) | 17313 (54.2%) | 18843 (54.0%) |
| **Male** | 1393 (47.7%) | 14634 (45.8%) | 16027 (46.0%) |
| **Race/Ethnicity** |  |  |  |
| **Hispanic** | 721 (24.7%) | 7844 (24.6%) | 8565 (24.6%) |
| **NHW** | 1355 (46.4%) | 13707 (42.9%) | 15062 (43.2%) |
| **NHB** | 330 (11.3%) | 5159 (16.1%) | 5489 (15.7%) |
| **AAPI** | 190 (6.5%) | 1637 (5.1%) | 1827 (5.2%) |
| **Multiple** | 45 (1.5%) | 396 (1.2%) | 441 (1.3%) |
| **Other/Unknown** | 282 (9.6%) | 3204 (10.0%) | 3486 (10.0%) |
| **Health System** |  |  |  |
| **A** | 58 (2.0%) | 1374 (4.3%) | 1432 (4.1%) |
| **B** | 312 (10.7%) | 1680 (5.3%) | 1992 (5.7%) |
| **C** | 43 (1.5%) | 700 (2.2%) | 743 (2.1%) |
| **D** | 8 (0.3%) | 265 (0.8%) | 273 (0.8%) |
| **E** | 189 (6.5%) | 2139 (6.7%) | 2328 (6.7%) |
| **F** | 113 (3.9%) | 1198 (3.8%) | 1311 (3.8%) |
| **G** | 43 (1.5%) | 422 (1.3%) | 465 (1.3%) |
| **H** | 23 (0.8%) | 464 (1.5%) | 487 (1.4%) |
| **I** | 40 (1.4%) | 807 (2.5%) | 847 (2.4%) |
| **J** | 23 (0.8%) | 920 (2.9%) | 943 (2.7%) |
| **K** | 36 (1.2%) | 1294 (4.1%) | 1330 (3.8%) |
| **L** | 82 (2.8%) | 506 (1.6%) | 588 (1.7%) |
| **M** | 30 (1.0%) | 200 (0.6%) | 230 (0.7%) |
| **N** | 104 (3.6%) | 1440 (4.5%) | 1544 (4.4%) |
| **O** | 45 (1.5%) | 741 (2.3%) | 786 (2.3%) |
| **P** | 45 (1.5%) | 588 (1.8%) | 633 (1.8%) |
| **Q** | 817 (28.0%) | 6755 (21.1%) | 7572 (21.7%) |
| **R** | 46 (1.6%) | 1428 (4.5%) | 1474 (4.2%) |
| **S** | 21 (0.7%) | 228 (0.7%) | 249 (0.7%) |
| **T** | 174 (6.0%) | 2229 (7.0%) | 2403 (6.9%) |
| **U** | 293 (10.0%) | 1128 (3.5%) | 1421 (4.1%) |
| **V** | 13 (0.4%) | 66 (0.2%) | 79 (0.2%) |
| **W** | 87 (3.0%) | 567 (1.8%) | 654 (1.9%) |
| **X** | 43 (1.5%) | 838 (2.6%) | 881 (2.5%) |
| **Y** | 7 (0.2%) | 591 (1.9%) | 598 (1.7%) |
| **Z** | 10 (0.3%) | 43 (0.1%) | 53 (0.2%) |
| **AA** | 66 (2.3%) | 1597 (5.0%) | 1663 (4.8%) |
| **AB** | 122 (4.2%) | 1480 (4.6%) | 1602 (4.6%) |
| **AC** | 30 (1.0%) | 259 (0.8%) | 289 (0.8%) |
| **Cohort Entry Month** |  |  |  |
| **04/2022** | 86 (2.9%) | 1958 (6.1%) | 2044 (5.9%) |
| **05/2022** | 293 (10.0%) | 3596 (11.3%) | 3889 (11.2%) |
| **06/2022** | 227 (7.8%) | 2832 (8.9%) | 3059 (8.8%) |
| **07/2022** | 344 (11.8%) | 2948 (9.2%) | 3292 (9.4%) |
| **08/2022** | 283 (9.7%) | 3568 (11.2%) | 3851 (11.0%) |
| **09/2022** | 178 (6.1%) | 2681 (8.4%) | 2859 (8.2%) |
| **10/2022** | 92 (3.1%) | 1425 (4.5%) | 1517 (4.4%) |
| **11/2022** | 125 (4.3%) | 1549 (4.8%) | 1674 (4.8%) |
| **12/2022** | 231 (7.9%) | 1930 (6.0%) | 2161 (6.2%) |
| **01/2023** | 154 (5.3%) | 1837 (5.8%) | 1991 (5.7%) |
| **02/2023** | 179 (6.1%) | 1493 (4.7%) | 1672 (4.8%) |
| **03/2023** | 103 (3.5%) | 971 (3.0%) | 1074 (3.1%) |
| **04/2023** | 66 (2.3%) | 509 (1.6%) | 575 (1.6%) |
| **05/2023** | 56 (1.9%) | 431 (1.3%) | 487 (1.4%) |
| **06/2023** | 40 (1.4%) | 251 (0.8%) | 291 (0.8%) |
| **07/2023** | 56 (1.9%) | 337 (1.1%) | 393 (1.1%) |
| **08/2023** | 201 (6.9%) | 1394 (4.4%) | 1595 (4.6%) |
| **09/2023** | 117 (4.0%) | 1269 (4.0%) | 1386 (4.0%) |
| **10/2023** | 35 (1.2%) | 278 (0.9%) | 313 (0.9%) |
| **11/2023** | 25 (0.9%) | 326 (1.0%) | 351 (1.0%) |
| **12/2023** | 32 (1.1%) | 364 (1.1%) | 396 (1.1%) |
| **Obesity** |  |  |  |
| **Nonobese** | 780 (26.7%) | 10097 (31.6%) | 10877 (31.2%) |
| **Obese** | 2110 (72.2%) | 21062 (65.9%) | 23172 (66.5%) |
| **Unknown** | 33 (1.1%) | 788 (2.5%) | 821 (2.4%) |
| **Chronic Disease** |  |  |  |
| **No Chronic Disease** | 2262 (77.4%) | 26560 (83.1%) | 28822 (82.7%) |
| **Non-complex Chronic Disease** | 374 (12.8%) | 3565 (11.2%) | 3939 (11.3%) |
| **Complex Chronic Disease** | 287 (9.8%) | 1822 (5.7%) | 2109 (6.0%) |
| **Severity during acute COVID-19** |  |  |  |
| **Asymptomatic** | 733 (25.1%) | 8270 (25.9%) | 9003 (25.8%) |
| **Mild** | 2100 (71.8%) | 22402 (70.1%) | 24502 (70.3%) |
| **Moderate** | 83 (2.8%) | 969 (3.0%) | 1052 (3.0%) |
| **Severe** | 7 (0.2%) | 306 (1.0%) | 313 (0.9%) |

### Incidences of Acute COVID-19 Illness Outcome Events and Post-acute Phase Diagnoses

**Table 2** summarizes the absolute risks and incidences for acute COVID-19 illness and post-acute phase diagnoses for the treated compared to the control group patients before propensity score matching. During the acute phase, the treated group demonstrated a notably lower absolute risk of hospitalization (0.58% vs. 0.96%), outpatient visits (74.17% vs. 82.66%), and ED visits (1.81% vs. 2.28%) compared to the control group. Similarly, the absolute risk of moderate/severe acute illness was lower in the treated group (3.08% vs. 3.99%). The analysis also extended to sixteen acute-illness symptoms and conditions, where the treated group generally exhibited lower incidences across most symptoms, with notable reductions in diarrhea, vomiting, and arrhythmias compared to their untreated counterparts. For example, the incidence of diarrhea was 0.44% in the treated group versus 0.64% in the untreated group, and arrhythmias were reported at 0.86% versus 1.18%, respectively. For PASC diagnosis (U09.9), the absolute risks are similar in the treated group (0.21%) compared to the control group (0.20%).

**Table 2.** Absolute risks and number of events before PS matching (incidence in %).

| **Outcome Name** |  | **Overall**<br>**(N=34,870)** | **Treated**<br>**(N=2,923)** | **Not Treated**<br>**(N=31,947)** |
| --- | --- | --- | --- | --- |
| **Acute Phase** |  |  |  |  |
| **Healthcare Utilization** |  |  |  |  |
|  | Hospitalization | 0.93(323) | 0.58(17) | 0.96(306) |
|  | Outpatient visit | 81.94(28574) | 74.17(2168) | 82.66(26406) |
|  | ED visit | 2.24(781) | 1.81(53) | 2.28(728) |
| **Acute Illness Severity** |  |  |  |  |
|  | Moderate/Severe acute illness | 3.92(1365) | 3.08(90) | 3.99(1275) |
| **Symptoms and Conditions** |  |  |  |  |
|  | Diarrhea | 0.62(207) | 0.44(12) | 0.64(195) |
|  | Nausea | 0.98(321) | 0.66(18) | 1.08(303) |
|  | Vomiting | 1.06(343) | 0.90(24) | 1.07(319) |
|  | Abdominal pain | 1.34(396) | 0.98(24) | 1.37(372) |
|  | Arrhythmias | 1.15(377) | 0.86(23) | 1.18(354) |
|  | Cardiovascular signs and symptoms | 0.59(197) | 0.47(13) | 0.60(184) |
|  | Chest pain | 1.77(569) | 0.82(22) | 1.86(547) |
|  | Fatigue and malaise | 1.83(581) | 0.86(22) | 1.92(559) |
|  | Fever and chills | 6.80(2138) | 5.26(140) | 6.94(1998) |
|  | Fluid and electrolyte | 0.12(41) | 0.14(4) | 0.12(37) |
|  | Generalized pain | 1.65(527) | 1.19(31) | 1.69(496) |
|  | Headache | 3.18(930) | 1.77(43) | 3.31(887) |
|  | Mental health | 3.052(734) | 1.89(34) | 3.15(700) |
|  | Musculoskeletal | 2.09(560) | 1.30(28) | 2.16(532) |
|  | Respiratory signs and symptoms | 12.63(3222) | 7.93(170) | 13.07(3052) |
|  | Skin | 1.11(342) | 0.92(23) | 1.13(319) |
| **Post-acute Phase** |  |  |  |  |
| **U09.9** |  |  |  |  |
|  | PASC diagnoses (U09.9) | 0.20(70) | 0.21(6) | 0.20(64) |

### Adjusted Relative Risk of Acute and Post-acute COVID-19 Illness Outcomes

Our analysis focused on the effectiveness of nirmatrelvir-ritonavir treatment associated with various acute phase outcomes and the specific post-acute phase diagnoses after adjusting for confounding factors via propensity score matching. **Table 3** summarizes the absolute risks and incidences for acute COVID-19 illness and post-acute phase diagnoses for the treated compared to the control group patients after propensity score matching. The RRs for these outcomes, as detailed in **Figure 2**, show that nirmatrelvir-ritonavir treatment was significantly associated with reductions in the risk of several key acute phase healthcare utilization outcomes including hospitalization (RR 0.48; 95% confidence interval (CI) 0.29-0.80) and outpatient visits (RR 0.86; 95% CI 0.82-0.90). Additionally, the treatment was linked with a decreased risk of moderate to severe acute illness (RR 0.69; 95% CI 0.56-0.87).

**Table 3.** Absolute risks and number of events after PS matching (incidence in %).

| Outcome Name | Overall<br>(N=13,074) | Treated<br>(N=2,920) | Not Treated<br>(N=15,994) |
| --- | --- | --- | --- |
| **Acute Phase** |  |  |  |
| **Healthcare Utilization** |  |  |  |
| Hospitalization | 1.04(166) | 0.55(16) | 1.15(150) |
| Outpatient visit | 83.87(13414) | 74.18(2166) | 86.03(11248) |
| ED visit | 2.23(357) | 1.82(53) | 2.33(304) |
| **Acute Illness Severity** |  |  |  |
| Moderate/Severe acute illness | 4.20(671) | 3.08(90) | 4.44(581) |
| **Symptoms and Conditions** |  |  |  |
| Diarrhea | 0.66(98) | 0.44(12) | 0.70(86) |
| Nausea | 0.89(133) | 0.63(17) | 0.95(116) |
| Vomiting | 0.98(144) | 0.90(24) | 1.00(120) |
| Abdominal pain | 1.31(176) | 0.98(24) | 1.39(152) |
| Arrhythmias | 1.08(159) | 0.86(23) | 1.13(136) |
| Cardiovascular signs and symptoms | 0.64(96) | 0.48(13) | 0.67(83) |
| Chest pain | 1.80(266) | 0.82(22) | 2.02(244) |
| Fatigue and malaise | 1.58(223) | 0.86(22) | 1.74(201) |
| Fever and chills | 6.06(880) | 5.27(140) | 6.24(740) |
| Fluid and electrolyte | 0.18(28) | 0.14(4) | 0.19(24) |
| Generalized pain | 1.71(244) | 1.20(31) | 1.83(213) |
| Headache | 2.72(359) | 1.77(43) | 2.93(316) |
| Mental health | 3.10(311) | 1.89(34) | 3.36(277) |
| Musculoskeletal | 1.92(226) | 1.30(28) | 2.06(198) |
| Respiratory signs and symptoms | 12.14(1417) | 7.96(170) | 13.08(1247) |
| Skin | 0.99(135) | 0.93(23) | 1.00(112) |
| **Post-acute Phase** |  |  |  |
| **U09.9** |  |  |  |
| PASC diagnoses (U09.9) | 0.21(34) | 0.21(6) | 0.21(28) |

**Figure 2.**
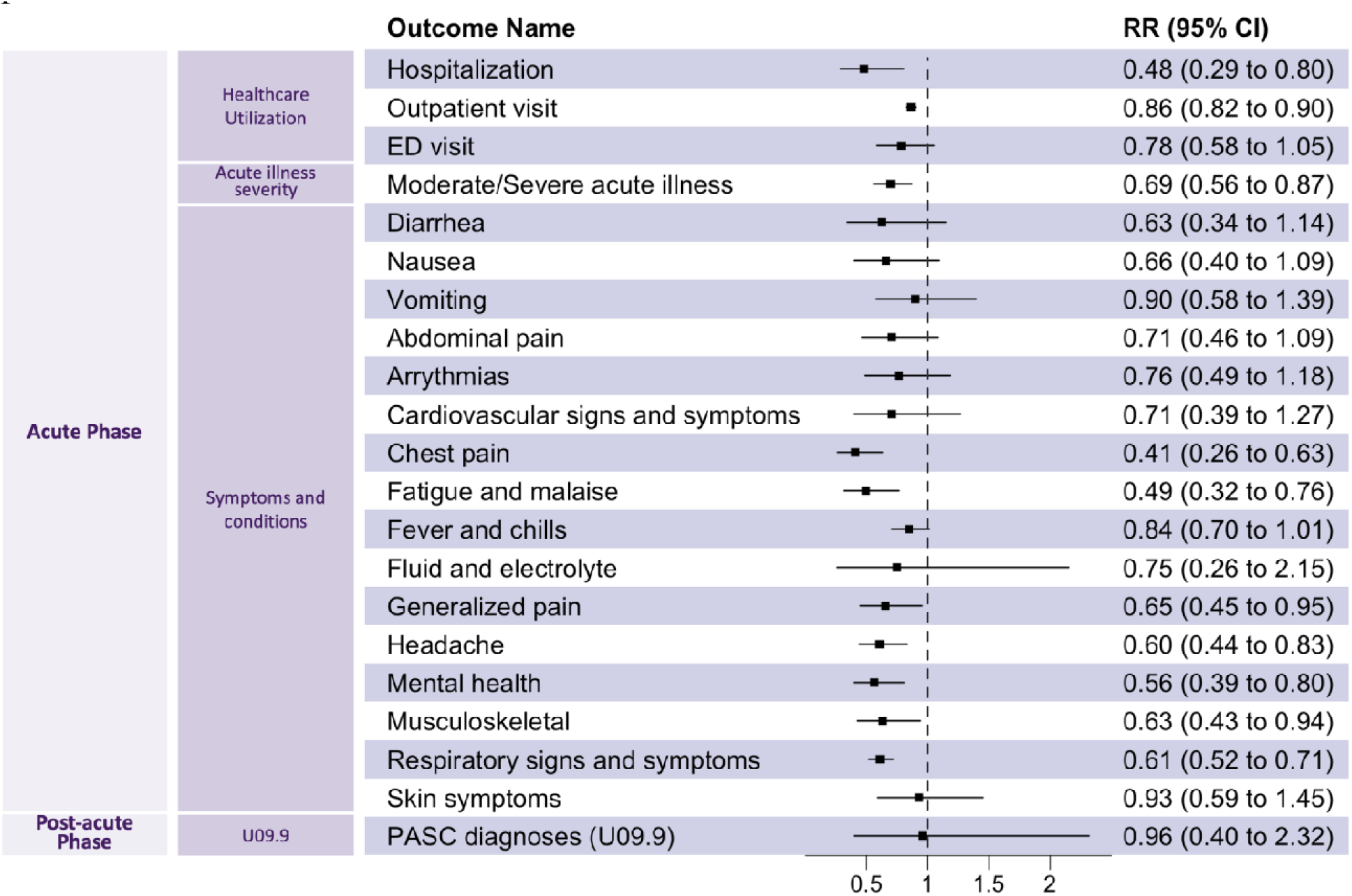
The relative risk (RR) of acute phase outcomes and post-acute phase diagnoses for patients treated and not treated with nirmatrelvir-ritonavir.

Acute COVID-19 illness symptoms and conditions also showed decreased risks in the treatment group compared with the control group. Notably, nirmatrelvir-ritonavir was associated with reduced risk of conditions in chest pain (RR 0.41; 95% CI 0.26-0.63), fatigue and malaise (RR 0.49; 95% CI 0.32-0.76), generalized pain (RR 0.65; 95% CI 0.45-0.95), headache (RR 0.60; 95% CI 0.44-0.83), mental health (RR 0.56; 95% CI 0.39-0.80), musculoskeletal (RR 0.63, 95% CI 0.43-0.94), respiratory signs and symptoms (RR 0.61, 95% CI 0.52-0.71). There was a lack of a statistically significant association between nirmatrelvir-ritonavir and other conditions, but showed a trend toward reducing diarrhea (RR 0.63; 95% CI 0.34-1.14), nausea (RR 0.66; 95% CI 0.40-1.09), vomiting (RR 0.90; 95% CI 0.58-1.39), abdominal pain (RR 0.71; 95% CI 0.46-1.09), arrhythmias (RR 0.76; 95% CI 0.49-1.18), cardiovascular signs and symptoms (RR 0.71; 95% CI 0.39-1.27), fever and chills (RR 0.84; 95% CI 0.7-1.01), fluid and electrolyte (RR 0.75; 95% CI 0.26-2.15) and skin symptoms (RR 0.93; 95% CI 0.59-1.45).

The RR for PASC diagnoses did not show a statistically significant reduction (RR 0.96; 95% CI 0.40-2.32).

### Sensitivity Analysis

In the sensitivity analysis, the inclusion of patients under 12 years old (Section S4) revealed that nirmatrelvir-ritonavir treatment was associated with reduced risks across several healthcare utilization outcomes and moderate/severe acute illness symptoms, like results seen in adolescents. And the impact on long COVID diagnoses was also modest and not statistically significant as the main analysis, with a slightly more moderate effect on generalized pain in this sensitivity analysis. Including adolescents with weights under 40 kg or missing data (Section S5) indicated a pronounced reduction in arrhythmias. Negative control experiments (Section S6) revealed a slight systematic error, evidenced by minor shifts in point estimates with wider CIs.

### Subgroup Analysis

In the subgroup analysis, adolescents aged 16 to 20 years showed a more pronounced reduction in healthcare utilization outcomes compared to those aged 12 to 15 years (Section S7), with significant decreases noted in chest pain, fatigue and malaise, headache, mental health, and respiratory symptoms. While the younger age group showed significant reductions in moderate/severe acute illness, chest pain, fatigue and malaise, mental health, respiratory signs and symptoms. Obese patients experienced more significant reductions in hospitalization visits and chest pain, while non-obese patients saw greater reductions in outpatient visits, moderate/severe acute illness, nausea, fatigue and malaise, fever and chills, headache, mental health, and respiratory signs and symptoms (Section S8). Vaccinated individuals demonstrated significant reductions across all measured healthcare utilization outcomes, including moderate/severe acute illness and a range of acute-phase symptoms among abdominal pain, chest pain, fatigue and malaise, fever and chills, headache, mental health, musculoskeletal, respiratory signs and symptoms and skin (Section S9). Patients with at least one risk factor for severe COVID-19 displayed slight reductions across most acute and post-acute outcomes, apart from hospitalization visits and chest pain, unlike those without any risk factors (Section S10). Lastly, patients who experienced severe COVID-19 during the acute phase exhibited significant reductions in hospitalization visits, outpatient visits, chest pain, fatigue and malaise, headache, and respiratory symptoms, whereas those in the non-severe group saw reductions primarily in outpatient and ED visits, mental health, and respiratory symptoms (Section S11).

## Discussion

In this study, we investigate the effectiveness of nirmatrelvir-ritonavir in managing both the acute and post-acute phases of SARS-CoV-2 infection among adolescents treated in outpatient settings. Our cohort analysis involved 2,923 adolescents who were treated with oral nirmatrelvir-ritonavir within 5 days after the positive test and not hospitalized on the day of the positive test result and 31,947 adolescents who did not receive any COVID-19 antiviral or antibody treatment and nirmatrelvir-ritonavir during the acute phase of SARS-CoV-2 infection.

Nirmatrelvir-ritonavir was found to be significantly associated with a reduced risk of hospitalization, outpatient visits, and moderate to severe illness during the acute phase. Additionally, nirmatrelvir-ritonavir appeared to be significantly associated with a reduced risk of a set of acute symptoms, including chest pain, fatigue and malaise, generalized pain, headache, mental health issues, musculoskeletal, and respiratory signs and symptoms. For the long COVID diagnoses, nirmatrelvir-ritonavir did not demonstrate a statistically significant effect in reducing risks. The effectiveness of nirmatrelvir-ritonavir was consistently observed across various subgroups, including different ages, obesity status, and the severity of acute COVID-19 symptoms. Notably, even adolescents without any risk factors for severe COVID-19 showed reductions in most acute and post-acute outcomes, highlighting its broad potential impact on those previously considered at low risk of acute COVID-19 illness. The robustness of our findings was affirmed through extensive sensitivity analyses and negative control experiments.

Our study had several strengths. Firstly, our study used the expansive RECOVER EHR database, which allowed us to construct a large-scale cohort comprising both COVID-exposed and non-exposed individuals, all of whom were followed longitudinally throughout the post-acute period. This is the largest study in the United States to date evaluating the effectiveness of nirmatrelvir-ritonavir in adolescents and encompasses a wide demographic cross-section of the U.S. pediatric population. This substantial cohort size of US adolescents enhances the robustness and comprehensiveness of our findings, making a significant contribution to understanding treatment impacts. Secondly, we used a propensity score matching method at a ratio of 1:5^33,34^, enhancing the balance between our comparative groups. This method, which handles numerous covariates, effectively accounts for the nonlinear influences of confounders, ensuring a more equitable and accurate cohort for our investigation^44^. Thirdly, in addition to examining both acute and post-acute phase outcomes, we conducted extensive sensitivity analyses to assess the robustness of our clinical findings. Additionally, we employed negative control experiments^42,43^ to calibrate systematic bias, controlling unmeasured confounders.

This study is subject to several limitations. Firstly, the selection of confounding variables for the propensity score model was informed by expert opinion and the availability of data. Despite this careful selection, the incompleteness of EHR data may still lead to the existence of some unmeasured confounders. There may still be residual confounding despite matching on many key patient- and site-level variables. To address this, we have employed negative control experiments in our sensitivity analysis, enhancing the reliability of our results. These controls were also chosen with expert guidance and based on data accessibility. Future studies could benefit from incorporating a broader range of negative control outcomes to further solidify findings. Secondly, the use of EHR data presents inherent risks of misclassification biases due to potential inaccuracies in recording SARS-CoV-2 infection status and inconsistencies in follow-up data. Furthermore, the widespread adoption of at-home rapid antigen tests later in the pandemic may have led to underreported testing frequencies within the EHR systems. To mitigate these biases, future studies could adopt established statistical methods^45,46^ to enhance data accuracy and reliability. Thirdly, potential existence of selection bias in the study cohort due to health-seeking behavior could affect the findings. Specifically, adolescents treated with nirmatrelvir-ritonavir might have been more likely to be symptomatic and might visit hospitals more frequently, potentially inflating the observed acute or post-acute outcomes. Despite that we have incorporated healthcare utilization factors like the number of inpatient, outpatient, ED visits, and negative COVID-19 tests as confounders, future analyses could benefit from considering additional variables such as socioeconomic status and health insurance coverage. Finally, the current databases present limitations due to the low number of events recorded for long COVID, which may result in limited statistical power and potential biases in our analysis. Here we utilized only the diagnosis code U09.9 to define long COVID outcome. We intend to address this limitation by conducting further studies as more patient data become available.

## Conclusions

This cohort study offers a comprehensive evaluation of the impact of nirmatrelvir-ritonavir treatment on adolescents diagnosed with SARS-CoV-2, focusing on its effect during both the acute and post-acute phases of COVID-19. Our findings demonstrate that taking nirmatrelvir-ritonavir within five days of a positive SARS-CoV-2 test is associated with a significant reduction in healthcare utilization, including lower rates of hospitalization and outpatient visits, as well as acute illness severity during the acute phase after the COVID-19 infection. Moreover, the treatment was effective in reducing risks of acute symptoms such as chest pain, fatigue and malaise, generalized pain, headache, mental health, musculoskeletal, and respiratory signs and symptoms, suggesting that nirmatrelvir-ritonavir not only mitigates rates of healthcare utilization and illness severity but may also alleviate certain acute symptoms.

However, when considering long COVID diagnosis, the benefits of nirmatrelvir-ritonavir appear more limited. Although there was a trend towards a reduction in the risk of PASC diagnosis U09.9, this finding did not reach statistical significance, indicating that the effectiveness of nirmatrelvir-ritonavir in preventing long-term sequelae is less certain. Our primary analysis, including adjustments for age and weight criteria, as well as empirical calibration using negative control outcomes, underscore the strength and reliability of these associations. This underscores the necessity for ongoing research to fully understand the potential of nirmatrelvir-ritonavir in managing the long-term consequences of COVID-19 among adolescents.

## Data availability

The results reported in this study are based on detailed individual-level patient data compiled as part of the RECOVER program. Due to the high risk of reidentification based on the number of unique patterns in the date, patient privacy regulations prohibit us from releasing the data publicly. The data are maintained in a secure enclave, with access managed by the program coordinating center to remain compliant with regulatory and program requirements. Please direct requests to access the data, either for the reproduction of the work reported here or for other purposes, to the RECOVER EHR Pediatric Coordinating Center.

## Code availability

The analysis code is available for interested readers by emailing the corresponding author, Dr. Yong Chen, through.

## Acknowledgments

This study is part of the NIH Researching COVID to Enhance Recovery (RECOVER) Initiative, which seeks to understand, treat, and prevent the long COVID. For more information on RECOVER, visit https://recovercovid.org/.

We would like to thank the National Community Engagement Group (NCEG), all patients, caregivers, and community Representatives, and all the participants enrolled in the RECOVER Initiative. We would like to thank the patient representatives Leyna Aragon and Etienne A. Carignan for their insights.

## References

1. Hammond, J. et al. Oral Nirmatrelvir for High-Risk, Nonhospitalized Adults with Covid-19. N. Engl. J. Med. 386, 1397–1408 (2022).

2. Hammond, J. et al. Nirmatrelvir for Vaccinated or Unvaccinated Adult Outpatients with Covid-19. N. Engl. J. Med. 390, 1186–1195 (2024).

3. Arbel, R. et al. Nirmatrelvir Use and Severe Covid-19 Outcomes during the Omicron Surge. N. Engl. J. Med. 387, 790–798 (2022).

4. Najjar-Debbiny, R. et al. Effectiveness of Paxlovid in Reducing Severe Coronavirus Disease 2019 and Mortality in High-Risk Patients. Clin. Infect. Dis. 76, E342–E349 (2023).

5. Dryden-Peterson, S. et al. Nirmatrelvir Plus Ritonavir for Early COVID-19 in a Large U.S. Health System : A Population-Based Cohort Study. Ann. Intern. Med. 176, 77–84 (2023).

6. Xie, Y., Choi, T. & Al-Aly, Z. Association of Treatment With Nirmatrelvir and the Risk of Post–COVID-19 Condition. JAMA Intern. Med. 183, 554–564 (2023).

7. Liu, T. H., Wu, J. Y., Huang, P. Y., Tsai, Y. W. & Lai, C. C. The effect of nirmatrelvir-ritonavir on the long-term risk of neuropsychiatric sequelae following COVID-19. J. Med. Virol. 95, (2023).

8. Liu, T. H., Wu, J. Y., Huang, P. Y., Tsai, Y. W. & Lai, C. C. The effect of nirmatrelvir plus ritonavir on the long-term risk of epilepsy and seizure following COVID-19: A retrospective cohort study including 91,528 patients. J. Infect. 86, 256–308 (2023).

9. Patel, R. et al. Incidence of Symptoms Associated with Post-Acute Sequelae of SARS-CoV-2 infection in Non-Hospitalized Vaccinated Patients Receiving Nirmatrelvir-Ritonavir. medRxiv 2023.04.05.23288196 (2023) doi:10.1101/2023.04.05.23288196.

10. Wang, H. et al. Association of nirmatrelvir–ritonavir with post-acute sequelae and mortality in patients admitted to hospital with COVID-19: a retrospective cohort study. Lancet Infect. Dis. 24, 1130–1140 (2024).

11. Wang, F. et al. Real-World Effectiveness of Nirmatrelvir in Protecting Long COVID for Outpatient Adult Patients – A Large-Scale Observational Cohort Study from the RECOVER Initiative. Res. Sq. (2024) doi:10.21203/RS.3.RS-4536807/V1.

12. Bajema, K. L. et al. Effectiveness of COVID-19 treatment with nirmatrelvir-ritonavir or molnupiravir among U.S. Veterans: target trial emulation studies with one-month and six-month outcomes. medRxiv 11, (2022).

13. Durstenfeld, M. S. et al. Association of nirmatrelvir for acute SARS-CoV-2 infection with subsequent Long COVID symptoms in an observational cohort study. J. Med. Virol. 96, (2024).

14. Congdon, S. et al. Nirmatrelvir/ritonavir and risk of long COVID symptoms: a retrospective cohort study. Sci. Reports 2023 131 13, 1–9 (2023).

15. Chuang, M. H. et al. Efficacy of nirmatrelvir and ritonavir for post-acute COVID-19 sequelae beyond 3 months of SARS-CoV-2 infection. J. Med. Virol. 95, (2023).

16. Geng, L. N. et al. Nirmatrelvir-Ritonavir and Symptoms in Adults With Postacute Sequelae of SARS-CoV-2 Infection: The STOP-PASC Randomized Clinical Trial. JAMA Intern. Med. 184, 1024–1034 (2024).

17. Management Strategies in Children and Adolescents with Mild to Moderate COVID-19. https://www.aap.org/en/pages/2019-novel-coronavirus-covid-19-infections/clinical-guidance/outpatient-covid-19-management-strategies-in-children-and-adolescents/.

18. FACT SHEET FOR HEALTHCARE PROVIDERS: EMERGENCY USE AUTHORIZATION FOR PAXLOVID. https://www.fda.gov/media/155050/download#:~:text=U.S. Food and Drug Administration,of pediatric patients 12 years.

19. CDC. Types of COVID-19 Treatment. https://www.cdc.gov/covid/treatment/index.html.

20. Bose-Brill, S. et al. Pediatric Nirmatrelvir/Ritonavir Prescribing Patterns During the COVID-19 Pandemic. Hosp. Pediatr. 14, e341–e348 (2024).

21. Yoshida, M. et al. Local and systemic responses to SARS-CoV-2 infection in children and adults. Nat. 2021 6027896 602, 321–327 (2021).

22. Brodin, P. Immune determinants of COVID-19 disease presentation and severity. Nat. Med. 2021 271 27, 28–33 (2021).

23. ClinicalTrials.gov. EPIC-Peds: A Study to Learn About the Study Medicine Called PF-07321332 (Nirmatrelvir)/Ritonavir in Patients Under 18 Years of Age With COVID-19 That Are Not Hospitalized But Are at Risk for Severe Disease. https://clinicaltrials.gov/study/NCT05261139.

24. Dalton, A. F. et al. 1937. Use of Paxlovid for Treatment of Acute COVID-19 and Occurrence of Post-COVID Conditions among Children and Adults at High-Risk for Severe COVID-19, April 1 - December 31, 2022. Open Forum Infect. Dis. 10, (2023).

25. Wong, C. K. H. et al. Effectiveness of nirmatrelvir/ritonavir in children and adolescents aged 12–17 years following SARS-CoV-2 Omicron infection: A target trial emulation. Nat. Commun. 15, 4917 (2024).

26. Hardin, A. P. et al. Age limit of pediatrics. Pediatrics 140, e20172151 (2017).

27. Infectious Diseases Society of America. Management of Drug Interactions With Nirmatrelvir/Ritonavir (Paxlovid®): Resource for Clinicians. https://www.idsociety.org/practice-guideline/covid-19-guideline-treatment-and-management/management-of-drug-interactions-with-nirmatrelvirritonavir-paxlovid/ (2022).

28. Razzaghi, H. et al. Vaccine Effectiveness Against Long COVID in Children. Pediatrics 153, 2023064446 (2024).

29. Skelly, A. C., Dettori, J. R. & Brodt, E. D. Assessing bias: the importance of considering confounding. Evid. Based. Spine. Care. J. 3, 9 (2012).

30. Wu, Q. et al. Real-world effectiveness of BNT162b2 against infection and severe diseases in children and adolescents. Ann. Intern. Med. 177, 165–176 (2024).

31. Simon, T. D., Haaland, W., Hawley, K., Lambka, K. & Mangione-Smith, R. Development and Validation of the Pediatric Medical Complexity Algorithm (PMCA) Version 3.0. Acad. Pediatr. 18, 577–580 (2018).

32. Rao, S. et al. Clinical Features and Burden of Postacute Sequelae of SARS-CoV-2 Infection in Children and Adolescents. JAMA Pediatr. 176, 1000–1009 (2022).

33. Woodward, M. Epidemiology: Study design and data analysis, third edition. Epidemiol. Study Des. Data Anal. Third Ed. 1–821 (2013) doi:10.1201/B16343/EPIDEMIOLOGY-MARK-WOODWARD/ACCESSIBILITY-INFORMATION.

34. Iwagami, M. & Shinozaki, T. Introduction to Matching in Case-Control and Cohort Studies. Ann. Clin. Epidemiol. 4, 33–40 (2022).

35. Austin, P. C. Balance diagnostics for comparing the distribution of baseline covariates between treatment groups in propensity-score matched samples. Stat. Med. 28, 3083 (2009).

36. Zou, G. A Modified Poisson Regression Approach to Prospective Studies with Binary Data. Am. J. Epidemiol. 159, 702–706 (2004).

37. Wu, Q., et al. Real-world effectiveness and causal mediation study of BNT162b2 on long COVID risks in children and adolescents. EClinicalMedicine 79, (2025).

38. Zhou, T. et al. Body mass index and postacute sequelae of SARS-CoV-2 infection in children and young adults. JAMA Netw. open 7, e2441970–e2441970 (2024).

39. Zhang, B., et al. Post-Acute Cardiovascular Outcomes of COVID-19 in Children and Adolescents: An EHR Cohort Study from the RECOVER Project. medRxiv (2024).

40. Forrest, C. B. et al. Severity of acute COVID-19 in children< 18 years old March 2020 to December 2021. Pediatrics 149, e2021055765 (2022).

41. Suchard, M. A. et al. Comprehensive comparative effectiveness and safety of first-line antihypertensive drug classes: a systematic, multinational, large-scale analysis. Lancet 394, 1816–1826 (2019).

42. Schuemie, M. J., Hripcsak, G., Ryan, P. B., Madigan, D. & Suchard, M. A. Empirical confidence interval calibration for population-level effect estimation studies in observational healthcare data. Proc. Natl. Acad. Sci. U. S. A. 115, 2571–2577 (2018).

43. Schuemie, M. J., Ryan, P. B., Dumouchel, W., Suchard, M. A. & Madigan, D. Interpreting observational studies: Why empirical calibration is needed to correct p-values. Stat. Med. 33, 209–218 (2014).

44. Amoah, J. et al. Comparing propensity score methods versus traditional regression analysis for the evaluation of observational data: a case study evaluating the treatment of gram-negative bloodstream infections. Clin. Infect. Dis. 71, e497–e505 (2020).

45. Chen, Y., Wang, J., Chubak, J. & Hubbard, R. A. Inflation of type I error rates due to differential misclassification in EHR-derived outcomes: Empirical illustration using breast cancer recurrence. Pharmacoepidemiol. Drug Saf. 28, 264–268 (2019).

46. Duan, R. et al. An empirical study for impacts of measurement errors on EHR based association studies. in AMIA Annual Symposium Proceedings vol. 2016 1764 (2017).

