## Supplementary Materials for "Assessment of Nirmatrelvir-Ritonavir Effects on Acute and Post-Acute COVID-19 Illness in US Adolescents: Target Trial Emulation"

### Section S1 Data Description

#### A. Description of electronic health records (EHR) data

The data used for our analysis is derived from electronic health records (EHRs), covering a wide range of healthcare interaction information routinely collected and stored by hospitals. This includes clinical data such as diagnoses and treatments, laboratory and test results, and administrative data including patient demographics and billing information. The hospital-based EHR data from the PEDSnet COVID-19 Database served as the basis for defining and determining exposure, outcomes, and covariates. In our study, we did not employ General Practitioner (GP) data, self-reported data, or any data sources external to the hospital’s EHR system. Compared with GP data, EHR data offers a more comprehensive and integrated view of patients’ health status, medical history, and healthcare interactions across different providers and settings. Our study strictly relies on structured, standardized EHR data entries made by healthcare providers within hospital settings.

#### B. Population and generalizability

The National Institutions of Health (NIH) launched the new RECOVER initiative in 2021 to leverage electronic health record (EHR) data to better identify and characterize patients with post-acute sequelae of SARS-CoV-2 infection (PASC). RECOVER obtains EHRs from three large national healthcare networks within the United States, covering regional catchment areas across 41 states. These networks collectively hold the EHRs of over 60 million patients, including records from more than 7 million individuals who have been affected by COVID-19. RECOVER collaborates with the National Institutes of Health's (NIH) All of Us Research Program, which contributes additional health records to this vast database. Together, these sources comprise one of the world's largest collections of EHRs.

In our study, participating institutions in this study included: Cincinnati Children’s Hospital Medical Center, Children’s Hospital of Philadelphia, Children’s Hospital of Colorado, Columbia University Irving Medical Center, Duke University, Intermountain Healthcare, University of Iowa Healthcare, University Medical Center New Orleans (LSU) – Institute for Public Health Innovation (IPHI), Ann & Robert H. Lurie Children’s Hospital of Chicago, Medical College of Wisconsin, University of Michigan, University of Missouri, Montefiore, Medical University of South Carolina, Nationwide Children’s Hospital, University of Nebraska Medical Center, Nemours Children’s Health System (in Delaware and Florida), Northwestern University, New York University School of Medicine, OCHIN, Inc., Ochsner Health System, Ohio State University, University of Pittsburgh/UPM, Stanford Children’s Health, Temple University, University of California, San Francisco, University of Florida/UF Health, University of Utah, UT Southwestern Medical Center, Vanderbilt University Medical Center, Wake Forest Baptist Health, and Weill Cornell Medical College. For this study, we used the s11 version of the data, collected till December 2023.

### Section S2 Supplemental Methods

#### A. Specification of hypothetical trials and the target trial emulation study

We detailed the hypothetical randomized controlled trials (RCT) we specified and the corresponding target trial emulation study to investigate the effectiveness of nirmatrelvir in reducing acute and post-acute COVID-19 illness in United States Adolescents in Table S1 below.

**Table S1.** Specification of hypothetical trials and the target trial emulation study.

| Protocol component | Hypothetical trial specification | Target Trial Emulation |
| --- | --- | --- |
| Eligibility criteria | - Aged 12-20 years at the time of positive SARS-CoV-2 test between April 01, 2022, and December 31, 2023 - Outpatient setting - Hospitalized at the day of cohort entry - Weight > 40kg - Exclude patients with diagnosis of CKD, ESRD, hepatic failure, hepatic insufficiency, or had procedure for chronic dialysis or kidney transplant - Exclude patients had contraindication medications up to 90 days before cohort entry | Similar to the hypothetical trial, patients without any interaction with the healthcare system during the baseline period were excluded to ensure they had an ongoing relationship with the health system. |
| Treatment strategies | (1) Receive Nirmatrelvir-ritonavir within 5 days after cohort entry  (2) Receive no Nirmatrelvir-ritonavir within 5 days after cohort entry | Same |
| Treatment assignment | Patients are randomly assigned to receive or not receive Nirmatrelvir-ritonavir | We assumed random assignment after propensity score matching using a list of confounders. |
| Outcomes | - COVID-19 acute illness severity during acute phase - Healthcare utilization (hospitalizations, ED visits, outpatient visits) during acute phase - 17 symptoms and conditions during acute phase | Same |
| Follow-up | For each person, follow-up began from the day of SARS-CoV-2 positive test and continued until study end | Same |
| Causal contrasts | Intention-to-treat (ITT) effect | Same |
| Statistical analysis | Modified Poisson regression | Same |

#### B. Study variables

**Table S2**. Variables used in the study evaluating the effectiveness of nirmatrelvir for adolescents concerning acute and post-acute COVID-19 illness.

| Variable | Functional form | Values | Detail | Codes/references |
| --- | --- | --- | --- | --- |
| Treatment (i.e., Exposure) |  |  |  |  |
| Treatment with nirmatrelvir based on prescription records | Indicator | Yes/No | A set of medication code including RxNorm, SPL, and NDC, based on records in the drug exposure domain. | See <https://github.com/nbxszby416/PASC-Nirmatrelvir> for detailed codes. |
| Outcome |  |  |  |  |
| Clinical PASC diagnosis | Indicator | Yes/No | Diagnosis code U09.9. | (1)in References |
| Visits to emergency department within 28 days after the entry | Indicator | Yes/No | Based on the condition occurrence and visit occurrence domains. |  |
| Hospitalization within 28 days after the entry | Indicator | Yes/No | Based on the condition occurrence and visit occurrence domains, including Inpatient Hospital Stay, Emergency Department Admit to Inpatient Hospital Stay, and Observation Stay |  |
| Outpatient visits within 28 days after the entry | Indicator | Yes/No | Based on the condition occurrence and visit occurrence domains including Ambulatory/Outpatient Visit (With a Physician) and Interactive Telemedicine Service |  |
| Stratification variables |  |  |  |  |
| Severity of COVID-19 during acute phase | 2 categories | Non-severe group  Severe group | The reference paper categorized severity during acute phase of COVID-19 into asymptomatic, mild, moderate, and severe. In this study, “non-severe” group including asymptomatic, “severe” group including mild, moderate and severe. | (2) in References |
| Confounding variables |  |  |  |  |
| Age (years) | Linear | NA | Based on records in the person domain. | Age is defined as the integer of (date – birth date)/365.25 |
| Sex | Indicator | Male/Female | Based on records in the person domain. |  |
| Race/Ethnicity | 6 categories | NHW  NHB  Hispanic  AAPI  Multiple  Other/unknown | Based on records in the person domain. |  |
| Obesity | 3 categories | Yes/No/Unknown | Based on records in the measurement domain.  If measured at age < 24*30.5 days, NHANES weight z score > 1.64  If measured at 24*30.5 < age <240*30.5, NHANES BMI z score > 1.64  If measured at age >= 240*30.5, BMI kg/m2 > 30 |  |
| PMCA (Pediatric Medical Complexity Algorithm) | 3 categories | No chronic condition (PMCA = 0)  Non-complex chronic condition (PMCA = 1)  Complex chronic condition comorbidities (PMCA = 2) | Based on the condition occurrence and visit occurrence domains. | (3) in References |
| Diagnosis of each chronic condition cluster in 24 months to 7 days prior to the entry | Indicator | Yes/No | 205 chronic condition clusters were defined based on the condition occurrence and visit occurrence domains. | (1) in References |
| Number of visits to emergency department in 24 months to 7 days prior to the entry | 4 categories | 0/1/2/≥3 | Based on the condition occurrence and visit occurrence domains. |  |
| Number of inpatient visits in 24 months to 7 days prior to the entry | 4 categories | 0/1/2/≥3 | Based on the condition occurrence and visit occurrence domains, including Inpatient Hospital Stay, Emergency Department Admit to Inpatient Hospital Stay, and Observation Stay. |  |
| Number of outpatient visits in 24 months to 7 days prior to the entry | 4 categories | 0/1/2/≥3 | Based on the condition occurrence and visit occurrence domains including Ambulatory/Outpatient Visit (With a Physician) and Interactive Telemedicine Service |  |
| Number of unique medications in 24 months to 7 days prior to the entry | 2 categories | ≤1/>1 | Based on the drug exposure domain. |  |
| Number of negative COVID-19 tests in 24 months to 7 days prior to the entry | 4 categories | 0/1/2/≥3 | Based on the observation derivation recover domain. |  |
| Number of COVID-19 vaccine doses prior to the entry | 4 categories | 0/1/2/≥3 | Based on the immunization domain, using either the presence of a CVX code designating an administered or patient-reported dose or of a source value containing the terms “COVID” or “sars” in the immunization table. | CVX code: 207, 208, 210, 211, 212, 213, 217, 218 |
| Interval since the last COVID-19 immunization date | 3 categories | No vaccination  <4 months  ≥4 months |  |  |
| Other variables for eligibility criteria |  |  |  |  |
| Prior encounter in 24 months to 7 days prior to the entry | Indicator | Yes/No | Based on the condition occurrence and visit occurrence domains. |  |
| Multisystem inflammatory syndrome in children (MIS-C) | Indicator | Yes/No | A set of diagnostic codes including ICD10CM, ICD10, ICD9CM, ICD9, and SNOMED. | See <https://github.com/nbxszby416/PASC-Nirmatrelvir> for detailed codes. |
| Chronic kidney disease (CKD) and End-stage renal disease (ESRD) | Indicator | Yes/No | A set of diagnostic codes including ICD10CM, ICD10, ICD9CM, ICD9, and SNOMED. | See <https://github.com/nbxszby416/PASC-Nirmatrelvir> for detailed codes. |
| Chronic dialysis and kidney transplant | Indicator | Yes/No | A set of procedure codes including CPT4, HCPCS, ICD10PCS, ICD9Proc, and SNOMED. | See <https://github.com/nbxszby416/PASC-Nirmatrelvir> for detailed codes. |
| Hepatic failure and hepatic insufficiency | Indicator | Yes/No | A set of diagnostic codes including ICD10CM, ICD10, and SNOMED. | See <https://github.com/nbxszby416/PASC-Nirmatrelvir> for detailed codes. |
| Contraindications | Indicator | Yes/No | A set of medication codes using RxNorm ingredient code. | See <https://github.com/nbxszby416/PASC-Nirmatrelvir> for detailed codes. |

Note: All the domains in the table above are based on the PEDSnet common data model (CDM). More details are available through this link: <https://data-models-service.research.chop.edu>.

### Section S3 Supplemental Results: patient characteristic balance between treated and control groups

#### A. Propensity-score (PS) models and matching

We fitted a large-scale PS model for the study cohort with baseline patient characteristics including

- Demographics (age at index date; sex; race/ethnicity)
- Obesity status
- Chronic condition indicator as defined by the Pediatric Medical Complexity Algorithm (PMCA)
  - No chronic condition (PMCA = 0)
  - Non-complex chronic condition (PMCA = 1)
  - Complex chronic condition comorbidities (PMCA = 2)
- The existence of a list of 205 chronic conditions 24 months ~ 7 days prior to the index
- Healthcare utilization 24 months ~ 7 days prior to index date categorized to 0,1,2, ≥3
  - Number of inpatient visits
  - Number of outpatient visits
  - Number of emergency department (ED) visits
  - Number of unique medications
  - Number of negative COVID-19 tests
- Vaccine information
  - Number of COVID-19 vaccine doses prior to the index, categorized to 0,1, ≥2
  - Interval since the last COVID-19 immunization date, categorized as no vaccine, <4 months, and ≥4 months
- Cohort entry date (index date) categorized to 1 month
- Healthcare system index

We exclude all covariates that occur in fewer than 10% of participants for computational efficiency. The treated group patients were then matched using the propensity score to control group patients with a ratio of 1:5 and caliper 0.2. The propensity score is estimated by the logistic model.

#### B. Empirical equipoise assessment

To assess the similarity across study groups, we present the preference score. This metric refines the propensity score by integrating treatment prevalence, facilitating an easily understood comparison. The preference score (F) is mathematically derived from the propensity score (S) and the treatment prevalence (P) using the following formula: (4,5)

$$\ln\left( \frac{F}{1-F} \right)=\ln\left( \frac{S}{1-S} \right)-\ln\left( \frac{P}{1-P} \right).$$

**Figure S1** below illustrates the preference score distributions for treatment and control groups, which indicates the high comparability of these studies.

**Figure S1**. Distribution of preference scores for treatment and control groups. Greater convergence of these distributions indicates a higher similarity in the predicted likelihood of receiving treatment between the comparison participants.

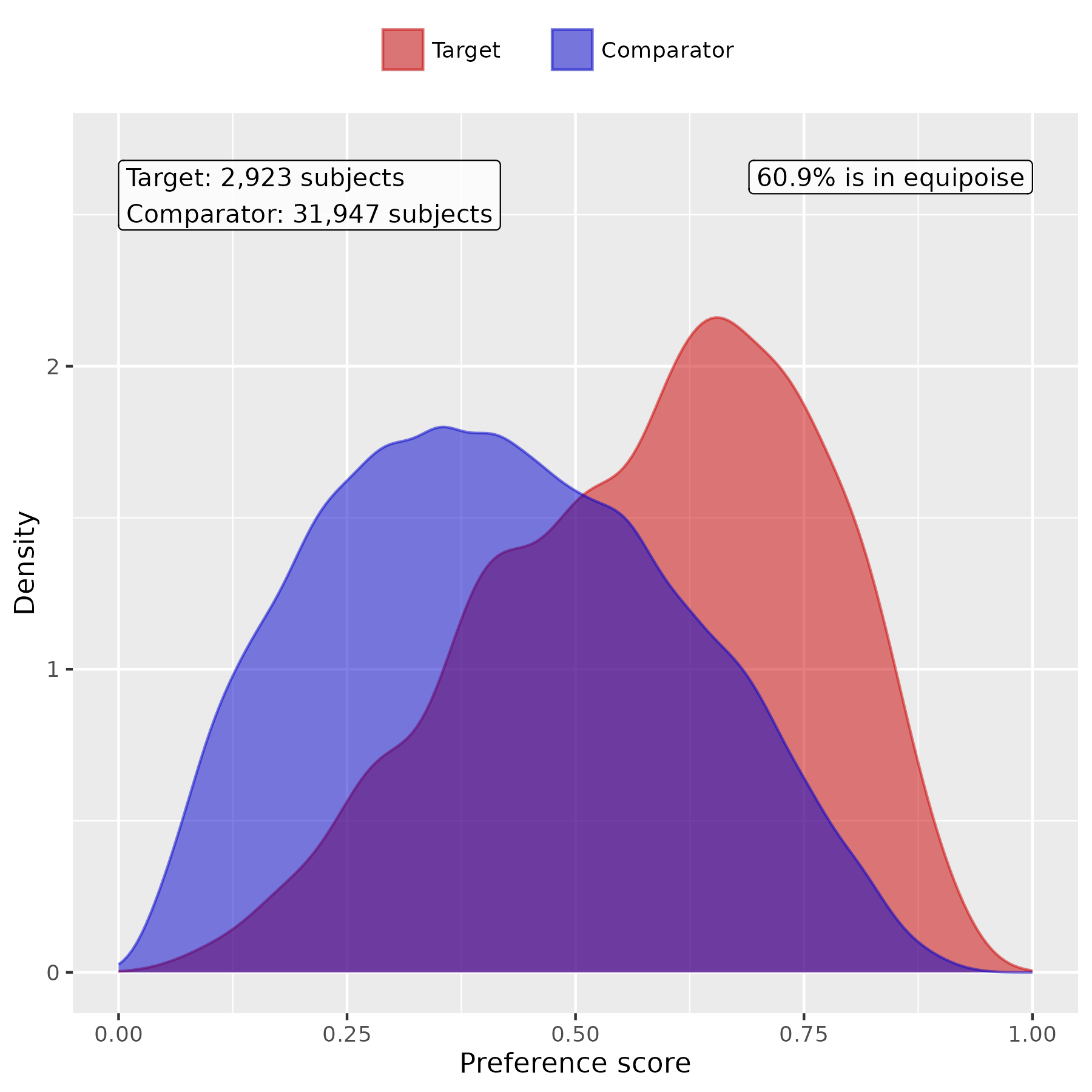

#### C. Patient characteristic balance in the primary analysis

We evaluate the balance of patient characteristics using the standardized mean difference (SMD). **Figure S2** presents the SMD of the study cohorts before and after PS score matching.

**Figure S2**. Patient characteristic balance before and after large-scale PS matching. The upper panel displays the top 20 covariates with the largest SMDs before matching, while the lower panel displays the top 20 covariates with the largest SMDs after matching.

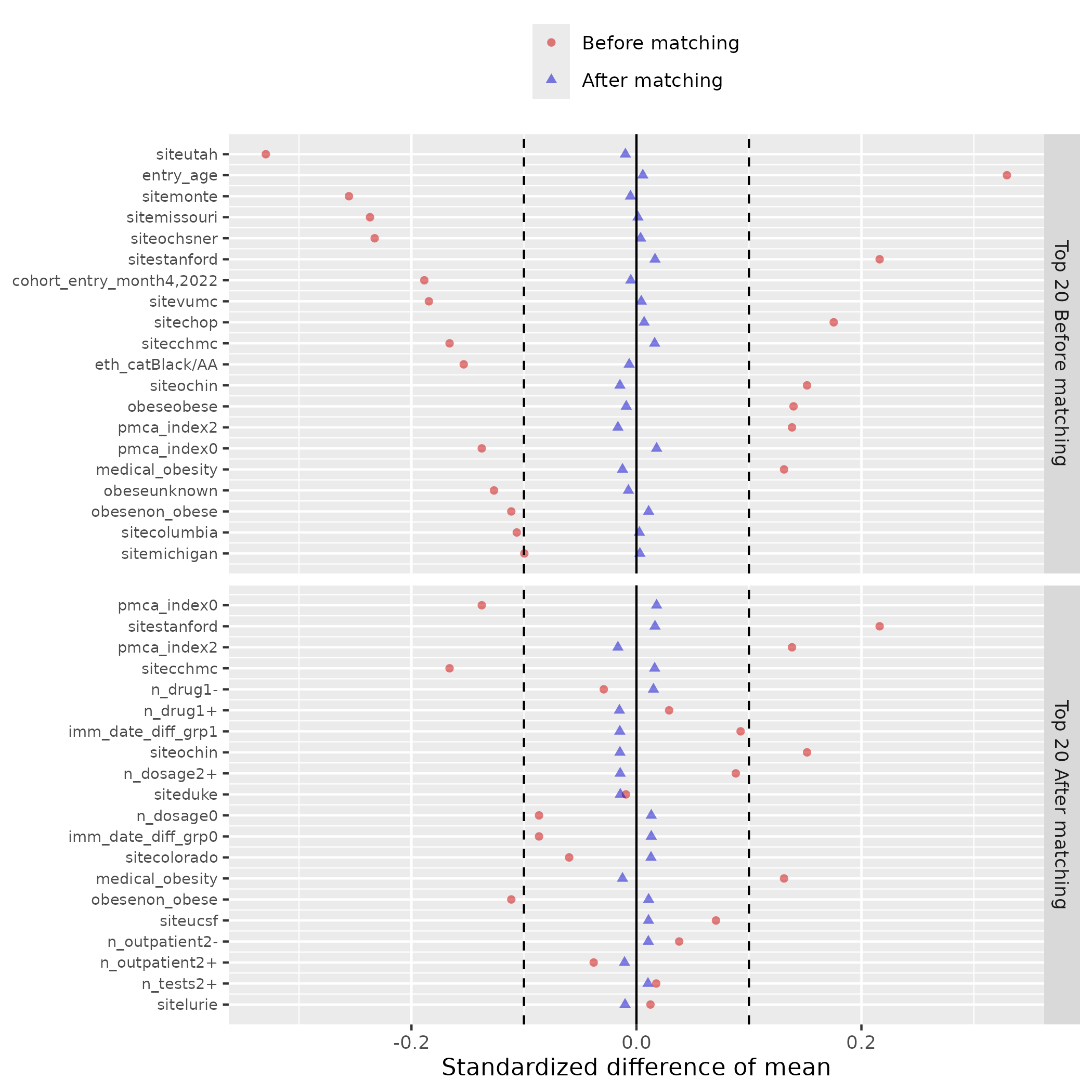

### Section S4 Supplemental Results: including patients aged under 12 at cohort entry

In the primary analysis, only adolescents aged between 12 and 21 are included. However, there can be off-label use of Nirmatrelvir for younger children aged under 12 although the medication is not approved for them. In this sensitivity analysis, we included both younger children aged under 12 and adolescents aged 12 to 21 at cohort entry.

**Table S3**. The relative risk (RR) of acute and post-acute outcomes for patients treated and not treated with Nirmatrelvir, including patients aged under 12.

| Outcome Name | RR (95% CI) |
| --- | --- |
| Hospitalization | **0.49(0.3-0.81)** |
| Outpatient visit | **0.86(0.82-0.90)** |
| ED visit | 0.80(0.59-1.07) |
| Moderate/Severe acute illness | **0.68(0.54-0.85)** |
| PASC diagnoses (U09.9) | 0.87(0.36-2.08) |
| diarrhea | 0.62(0.34-1.13) |
| nausea | 0.62(0.38-1.02) |
| vomiting | 0.93(0.60-1.45) |
| abdominal pain | 0.70(0.45-1.07) |
| arrythmias | 0.79(0.51-1.23) |
| cardiovascular signs and symptoms | 0.64(0.36-1.15) |
| chest pain | **0.44(0.28-0.67)** |
| fatigue and malaise | **0.48(0.31-0.74)** |
| fever and chills | 0.86(0.72-1.03) |
| fluid and electrolyte | 0.99(0.34-2.93) |
| generalized pain | 0.69(0.47-1.01) |
| headache | **0.56(0.41-0.78)** |
| mental health | **0.57(0.40-0.81)** |
| musculoskeletal | **0.67(0.45-0.99)** |
| respiratory signs and symptoms | **0.62(0.53-0.73)** |
| Skin symptoms | 0.98(0.62-1.53) |

### Section S5 Supplemental Results: including patients weighing under 40kg

In the primary analysis, only adolescents weighing more than 40kg are included. However, there can be off-label use of Nirmatrelvir for adolescents weighing less than 40kg although the medication is not approved for them. In this sensitivity analysis, we included adolescents aged 12 to 21 regardless of whether their weights are more or less than 40kg.

**Table S4**. The relative risk (RR) of acute and post-acute outcomes for patients treated and not treated with Nirmatrelvir, including patients weighing under 40 kg.

| Outcome Name | RR (95% CI) |
| --- | --- |
| Hospitalization | **0.52(0.32-0.83)** |
| Outpatient visit | **0.88(0.84-0.92)** |
| ED visit | 0.87(0.66-1.16) |
| Moderate/Severe acute illness | **0.72(0.58-0.90)** |
| PASC diagnoses (U09.9) | 0.83(0.35-1.97) |
| diarrhea | 0.65(0.36-1.19) |
| nausea | 0.72(0.44-1.17) |
| vomiting | 0.90(0.58-1.40) |
| abdominal pain | 0.70(0.46-1.07) |
| arrythmias | **0.63(0.41-0.97)** |
| cardiovascular signs and symptoms | 0.69(0.39-1.21) |
| chest pain | **0.41(0.27-0.62)** |
| fatigue and malaise | **0.48(0.32-0.73)** |
| fever and chills | 0.88(0.74-1.04) |
| fluid and electrolyte | 0.86(0.30-2.52) |
| generalized pain | **0.69(0.48-0.99)** |
| headache | **0.61(0.45-0.82)** |
| mental health | **0.55(0.39-0.77)** |
| musculoskeletal | **0.64(0.44-0.94)** |
| respiratory signs and symptoms | **0.66(0.57-0.76)** |
| Skin symptoms | 0.75(0.49-1.15) |

### Section S6 Supplemental Results: negative control experiments

To assess the robustness of our method, we conducted negative control experiments using a set of 36 negative control outcomes. Negative control outcomes are clinical outcomes believed to have no causal relationship with the treatment. The list of outcomes was carefully selected by pediatric physicians and includes: acne, astigmatism, autism/autistic disorder, closed fracture of distal end of radius, closed injury of head, concussion, contact dermatitis, diaper rash, displacements – bone, epilepsy, falls, foreign body in ear, impetigo, inguinal hernia, injury of finger, injury of free lower limb, injury of head, injury of left leg, injury of right foot, injury of right hand, injury of right leg, injury of upper extremity, insect bite, myopia, plagiocephaly, scoliosis, seizure, snoring/obstructive sleep apnea, speech delay, speech dysfunction, sprain of ankle, tinea capitis, tinea corporis, tongue tie, umbilical hernia, and wax in ear/impacted cerumen.(6–12)

We began by calculating the empirical null distribution of the negative control outcomes, which was subsequently used to adjust the RRs in the primary analysis. The methodology for determining the empirical null distribution and performing adjustments is thoroughly detailed in the works of Schuemie et al., published in 2014 and 2018.(13,14) These adjustments enhance the reliability of our findings by accounting for potential biases and ensuring robust estimation.

**Table S5**. Risks of acute-phase and post-acute phase COVID-19 outcomes, after negative control outcomes calibration.

| Outcome Name | RR (95% CI) |
| --- | --- |
| Hospitalization | 0.48(0.29-0.80) |
| Outpatient visit | **0.86(0.82-0.90)** |
| ED visit | 0.78(0.58-1.05) |
| Moderate/Severe acute illness | **0.69(0.56-0.87)** |
| PASC diagnoses (U09.9) | 0.96(0.40-2.32) |
| diarrhea | 0.63(0.34-1.14) |
| nausea | 0.66(0.40-1.09) |
| vomiting | 0.90(0.58-1.39) |
| abdominal pain | 0.71(0.46-1.09) |
| arrythmias | 0.76(0.49-1.18) |
| cardiovascular signs and symptoms | 0.71(0.39-1.27) |
| chest pain | **0.41(0.26-0.63)** |
| fatigue and malaise | **0.49(0.32-0.76)** |
| fever and chills | 0.84(0.70-1.01) |
| fluid and electrolyte | 0.75(0.26-2.15) |
| generalized pain | **0.65(0.45-0.95)** |
| headache | **0.60(0.44-0.83)** |
| mental health | **0.56(0.39-0.80)** |
| musculoskeletal | **0.63(0.43-0.94)** |
| respiratory signs and symptoms | **0.61(0.52-0.71)** |
| Skin symptoms | 0.93(0.59-1.45) |

### Section S7 Supplemental Results: stratified analysis by age subgroups

We conducted a subgroup analysis by the age group (12-15 years and 16-20 years).

**Table S6**. The relative risk (RR) of acute and post-acute outcomes for patients treated and not treated with Nirmatrelvir, patients aged 12 to 15 years.

| Outcome Name | RR (95% CI) |
| --- | --- |
| Hospitalization | 0.39(0.14-1.08) |
| Outpatient visit | **0.85(0.78-0.92)** |
| ED visit | 0.55(0.3-1.03) |
| Moderate/Severe acute illness | **0.47(0.3-0.76)** |
| PASC diagnoses (U09.9) | 2.87(0.69-12.01) |
| diarrhea | 0.24(0.03-1.79) |
| nausea | 0.73(0.28-1.86) |
| vomiting | 0.80(0.36-1.79) |
| abdominal pain | 0.77(0.38-1.55) |
| arrythmias | 0.57(0.22-1.44) |
| cardiovascular signs and symptoms | 0.14(0.02-1.01) |
| chest pain | **0.07(0.01-0.48)** |
| fatigue and malaise | **0.35(0.13-0.96)** |
| fever and chills | 0.88(0.65-1.19) |
| fluid and electrolyte | - |
| generalized pain | 0.90(0.46-1.77) |
| headache | 0.58(0.33-1.04) |
| mental health | **0.41(0.18-0.95)** |
| musculoskeletal | 0.54(0.25-1.18) |
| respiratory signs and symptoms | **0.64(0.48-0.86)** |
| Skin symptoms | 0.76(0.34-1.68) |

**Table S7.** The relative risk (RR) of acute and post-acute outcomes for patients treated and not treated with Nirmatrelvir, patients aged 16 to 20 years.

| Outcome Name | RR (95% CI) |
| --- | --- |
| Hospitalization | **0.53(0.30-0.94)** |
| Outpatient visit | **0.87(0.83-0.92)** |
| ED visit | 0.93(0.67-1.31) |
| Moderate/Severe acute illness | 0.80(0.62-1.03) |
| PASC diagnoses (U09.9) | 0.51(0.15-1.70) |
| diarrhea | 0.86(0.45-1.64) |
| nausea | 0.62(0.35-1.12) |
| vomiting | 0.95(0.56-1.61) |
| abdominal pain | 0.63(0.37-1.08) |
| arrythmias | 0.83(0.50-1.37) |
| cardiovascular signs and symptoms | 0.94(0.50-1.75) |
| chest pain | **0.59(0.38-0.94)** |
| fatigue and malaise | **0.52(0.32-0.85)** |
| fever and chills | 0.83(0.66-1.03) |
| fluid and electrolyte | 1.14(0.38-3.43) |
| generalized pain | 0.64(0.41-1.01) |
| headache | **0.6(0.41-0.87)** |
| mental health | **0.63(0.42-0.94)** |
| musculoskeletal | 0.66(0.41-1.04) |
| respiratory signs and symptoms | **0.61(0.50-0.74)** |
| Skin symptoms | 0.75(0.44-1.27) |

### Section S8 Supplemental Results: stratified analysis by obese status

We conducted a subgroup analysis by the obese status.

**Table S8**. The relative risk (RR) of acute and post-acute outcomes for patients treated and not treated with Nirmatrelvir, patients with obese.

| Outcome Name | RR (95% CI) |
| --- | --- |
| Hospitalization | **0.47(0.24-0.94)** |
| Outpatient visit | **0.92(0.87-0.97)** |
| ED visit | 0.88(0.61-1.25) |
| Moderate/Severe acute illness | **0.71(0.53-0.94)** |
| PASC diagnoses (U09.9) | 0.78(0.23-2.67) |
| diarrhea | 0.61(0.31-1.23) |
| nausea | 0.88(0.52-1.47) |
| vomiting | 0.85(0.51-1.41) |
| abdominal pain | 0.89(0.53-1.47) |
| arrythmias | 0.96(0.56-1.65) |
| cardiovascular signs and symptoms | 0.91(0.47-1.74) |
| chest pain | **0.40(0.23-0.71)** |
| fatigue and malaise | **0.59(0.36-0.96)** |
| fever and chills | 0.89(0.73-1.09) |
| fluid and electrolyte | 0.99(0.21-4.56) |
| generalized pain | 0.65(0.42-1.00) |
| headache | 0.75(0.53-1.06) |
| mental health | 0.71(0.47-1.07) |
| musculoskeletal | 0.69(0.44-1.08) |
| respiratory signs and symptoms | **0.69(0.58-0.82)** |
| Skin symptoms | 1.04(0.64-1.69) |

**Table S9**. The relative risk (RR) of acute and post-acute outcomes for patients treated and not treated with Nirmatrelvir, patients without obese.

| Outcome Name | RR (95% CI) |
| --- | --- |
| Hospitalization | 0.52(0.24-1.14) |
| Outpatient visit | **0.69(0.63-0.77)** |
| ED visit | 0.78(0.47-1.31) |
| Moderate/Severe acute illness | **0.67(0.47-0.95)** |
| PASC diagnoses (U09.9) | 1.03(0.29-3.60) |
| diarrhea | 0.75(0.22-2.55) |
| nausea | **0.11(0.02-0.83)** |
| vomiting | 1.07(0.44-2.6) |
| abdominal pain | 0.52(0.22-1.22) |
| arrythmias | 0.60(0.27-1.32) |
| cardiovascular signs and symptoms | 0.49(0.11-2.12) |
| chest pain | **0.46(0.23-0.92)** |
| fatigue and malaise | **0.27(0.1-0.73)** |
| fever and chills | **0.66(0.44-0.99)** |
| fluid and electrolyte | 1.47(0.30-7.30) |
| generalized pain | 0.71(0.34-1.50) |
| headache | **0.24(0.10-0.58)** |
| mental health | **0.29(0.14-0.63)** |
| musculoskeletal | 0.44(0.19-1.03) |
| respiratory signs and symptoms | **0.38(0.26-0.56)** |
| Skin symptoms | 0.36(0.11-1.16) |

### Section S9 Supplemental Results: stratified analysis by COVID-19 vaccine status

We conducted a subgroup analysis by the COVID-19 vaccine status.

**Table S10**. The relative risk (RR) of acute and post-acute outcomes for patients treated and not treated with Nirmatrelvir, patients receiving at least one COVID-19 vaccine dosage.

| Outcome Name | RR (95% CI) |
| --- | --- |
| Hospitalization | 2.04(0.71-5.89) |
| Outpatient visit | 1.00(0.93-1.08) |
| ED visit | 1.26(0.66-2.38) |
| Moderate/Severe acute illness | 0.82(0.46-1.45) |
| PASC diagnoses (U09.9) | 0.75(0.17-3.35) |
| diarrhea | 1.03(0.39-2.71) |
| nausea | 0.93(0.41-2.09) |
| vomiting | 1.66(0.83-3.32) |
| abdominal pain | 1.41(0.72-2.78) |
| arrythmias | 1.04(0.46-2.38) |
| cardiovascular signs and symptoms | 0.88(0.39-1.97) |
| chest pain | 0.62(0.28-1.36) |
| fatigue and malaise | 0.61(0.28-1.35) |
| fever and chills | 0.88(0.61-1.27) |
| fluid and electrolyte | - |
| generalized pain | 0.6(0.29-1.25) |
| headache | 0.57(0.31-1.04) |
| mental health | 0.90(0.53-1.54) |
| musculoskeletal | 0.98(0.51-1.87) |
| respiratory signs and symptoms | **0.74(0.57-0.97)** |
| Skin symptoms | 1.66(0.90-3.06) |

**Table S11**. The relative risk (RR) of acute and post-acute outcomes for patients treated and not treated with Nirmatrelvir, patients receiving no COVID-19 vaccine dosage.

| Outcome Name | RR (95% CI) |
| --- | --- |
| Hospitalization | **0.39(0.22-0.70)** |
| Outpatient visit | **0.79(0.74-0.84)** |
| ED visit | **0.69(0.49-0.96)** |
| Moderate/Severe acute illness | **0.67(0.53-0.85)** |
| PASC diagnoses (U09.9) | 0.67(0.24-1.93) |
| diarrhea | 0.48(0.22-1.05) |
| nausea | 0.54(0.29-1.02) |
| vomiting | 0.61(0.34-1.09) |
| abdominal pain | **0.52(0.29-0.93)** |
| arrythmias | 0.80(0.47-1.35) |
| cardiovascular signs and symptoms | 0.47(0.20-1.09) |
| chest pain | **0.38(0.22-0.64)** |
| fatigue and malaise | **0.41(0.24-0.69)** |
| fever and chills | **0.79(0.64-0.97)** |
| fluid and electrolyte | 1.25(0.41-3.80) |
| generalized pain | 0.79(0.51-1.24) |
| headache | **0.54(0.37-0.78)** |
| mental health | **0.46(0.28-0.74)** |
| musculoskeletal | **0.53(0.32-0.87)** |
| respiratory signs and symptoms | **0.56(0.46-0.68)** |
| Skin symptoms | **0.44(0.22-0.87)** |

### Section S10 Supplemental Results: stratified analysis by risk factor status

We conducted a subgroup analysis by the risk factor status. The risk factors include obesity, cardiovascular disease, asthma, hypertension, diabetes, neurological disease, genetic/metabolic disease, and cancer.

**Table S12**. The relative risk (RR) of acute and post-acute outcomes for patients treated and not treated with Nirmatrelvir, patients with at least one risk factor.

| Outcome Name | RR (95% CI) |
| --- | --- |
| Hospitalization | **0.45(0.23-0.89)** |
| Outpatient visit | **0.92(0.87-0.97)** |
| ED visit | 0.82(0.58-1.18) |
| Moderate/Severe acute illness | **0.72(0.55-0.96)** |
| PASC diagnoses (U09.9) | 0.74(0.22-2.51) |
| diarrhea | 0.69(0.34-1.39) |
| nausea | 0.96(0.56-1.65) |
| vomiting | 0.87(0.53-1.44) |
| abdominal pain | 0.91(0.55-1.51) |
| arrythmias | 0.80(0.47-1.36) |
| cardiovascular signs and symptoms | 0.71(0.38-1.35) |
| chest pain | **0.41(0.23-0.73)** |
| fatigue and malaise | **0.61(0.37-0.99)** |
| fever and chills | 0.92(0.75-1.12) |
| fluid and electrolyte | 0.81(0.18-3.64) |
| generalized pain | 0.69(0.44-1.07) |
| headache | 0.73(0.52-1.02) |
| mental health | 0.74(0.49-1.13) |
| musculoskeletal | 0.68(0.44-1.07) |
| respiratory signs and symptoms | **0.70(0.59-0.84)** |
| Skin symptoms | 1.08(0.66-1.75) |

**Table S13**. The relative risk (RR) of acute and post-acute outcomes for patients treated and not treated with Nirmatrelvir, patients without any risk factor.

| Outcome Name | RR (95% CI) |
| --- | --- |
| Hospitalization | 0.58(0.26-1.27) |
| Outpatient visit | **0.70(0.63-0.77)** |
| ED visit | 0.74(0.44-1.23) |
| Moderate/Severe acute illness | **0.69(0.48-0.98)** |
| PASC diagnoses (U09.9) | 0.89(0.26-3.08) |
| diarrhea | 0.75(0.22-2.56) |
| nausea | **0.13(0.02-0.96)** |
| vomiting | 1.11(0.45-2.72) |
| abdominal pain | 0.54(0.23-1.27) |
| arrythmias | 0.59(0.27-1.29) |
| cardiovascular signs and symptoms | 0.52(0.12-2.26) |
| chest pain | **0.48(0.24-0.95)** |
| fatigue and malaise | **0.27(0.10-0.74)** |
| fever and chills | **0.61(0.41-0.91)** |
| fluid and electrolyte | 1.26(0.26-6.08) |
| generalized pain | 0.74(0.35-1.57) |
| headache | **0.2(0.07-0.56)** |
| mental health | **0.28(0.13-0.6)** |
| musculoskeletal | **0.4(0.17-0.92)** |
| respiratory signs and symptoms | **0.36(0.24-0.54)** |
| Skin symptoms | 0.31(0.10-1.00) |

### Section S11 Supplemental Results: stratified analysis by severity of acute phase COVID-19 subgroups

We conducted a subgroup analysis by the severity of acute phase COVID-19. The severity of COVID-19 at the index date was stratified into the following four levels: asymptomatic, mild (symptomatic but without severe complications), moderate (characterized by more serious conditions associated with COVID-19, such as gastroenteritis, dehydration, and pneumonia), and severe (marked by critical conditions necessitating ICU care or mechanical ventilation)(2). For this sensitivity analysis, we grouped individuals presenting with asymptomatic symptoms into a “non-severe” category, while categorizing all remaining cases under a “severe” group.

**Table S14**. The relative risk (RR) of acute and post-acute outcomes for patients treated and not treated with Nirmatrelvir, within in the severe group.

| Outcome Name | RR (95% CI) |
| --- | --- |
| Hospitalization | **0.54(0.30-0.96)** |
| Outpatient visit | **0.95(0.90-0.99)** |
| ED visit | 1.01(0.74-1.36) |
| PASC diagnoses (U09.9) | 0.79(0.27-2.29) |
| diarrhea | 0.62(0.33-1.17) |
| nausea | 0.74(0.45-1.22) |
| vomiting | 0.83(0.54-1.28) |
| abdominal pain | 0.83(0.54-1.28) |
| arrythmias | 1.03(0.65-1.61) |
| cardiovascular signs and symptoms | 0.74(0.39-1.41) |
| chest pain | **0.55(0.36-0.86)** |
| fatigue and malaise | **0.55(0.35-0.86)** |
| fever and chills | 0.83(0.69-1.00) |
| fluid and electrolyte | 1.14(0.38-3.40) |
| generalized pain | 0.83(0.56-1.21) |
| headache | **0.57(0.41-0.79)** |
| mental health | 0.75(0.51-1.10) |
| musculoskeletal | 0.75(0.49-1.14) |
| respiratory signs and symptoms | **0.62(0.53-0.73)** |
| Skin symptoms | 1.06(0.66-1.70) |

**Table S15**. The relative risk (RR) of acute and post-acute outcomes for patients treated and not treated with Nirmatrelvir, within in the non-severe group.

| Outcome Name | RR (95% CI) |
| --- | --- |
| Hospitalization | 0.72(0.24-2.15) |
| Outpatient visit | **0.41(0.35-0.48)** |
| ED visit | **0.21(0.05-0.90)** |
| PASC diagnoses (U09.9) | 0.96(0.19-4.75) |
| diarrhea | 0.42(0.05-3.42) |
| nausea | - |
| vomiting | - |
| abdominal pain | - |
| arrythmias | - |
| cardiovascular signs and symptoms | 0.29(0.07-1.23) |
| chest pain | - |
| fatigue and malaise | - |
| fever and chills | - |
| fluid and electrolyte | - |
| generalized pain | - |
| headache | 0.58(0.13-2.63) |
| mental health | **0.14(0.04-0.44)** |
| musculoskeletal | 0.31(0.09-1.01) |
| respiratory signs and symptoms | **0.16(0.04-0.68)** |
| Skin sypmtoms | 0.24(0.06-1.01) |

### Reference

1.	Rao S, Lee GM, Razzaghi H, Lorman V, Mejias A, Pajor NM, et al. Clinical features and burden of postacute sequelae of SARS-CoV-2 infection in children and adolescents. JAMA Pediatr. 2022;176(10):1000–9.

2.	Forrest CB, Burrows EK, Mejias A, Razzaghi H, Christakis D, Jhaveri R, et al. Severity of acute COVID-19 in children< 18 years old March 2020 to December 2021. Pediatrics. 2022;149(4):e2021055765.

3.	Simon TD, Haaland W, Hawley K, Lambka K, Mangione-Smith R. Development and validation of the Pediatric Medical Complexity Algorithm (PMCA) version 3.0. Acad Pediatr. 2018;18(5):577–80.

4.	Walker AM, Patrick AR, Lauer MS, Hornbrook MC, Marin MG, Platt R, et al. A tool for assessing the feasibility of comparative effectiveness research. Comparative effectiveness research. 2013;11–20.

5.	Chapter 12 Population-Level Estimation | The Book of OHDSI. [Internet]. Available from: https://ohdsi.github.io/TheBookOfOhdsi/PopulationLevelEstimation.html

6.	Wu Q, Tong J, Zhang B, Zhang D, Chen J, Lei Y, et al. Real-World Effectiveness of BNT162b2 Against Infection and Severe Diseases in Children and Adolescents. Ann Intern Med. 2024;

7.	Wu Q, Zhang B, Tong J, Bailey LC, Bunnell HT, Chen J, et al. Real-world effectiveness and causal mediation study of BNT162b2 on long COVID risks in children and adolescents. EClinicalMedicine [Internet]. 2025;79:102962. Available from: https://www.sciencedirect.com/science/article/pii/S2589537024005418

8.	Zhou T, Zhang B, Zhang D, Wu Q, Chen J, Li L, et al. Body Mass Index and Postacute Sequelae of SARS-CoV-2 Infection in Children and Young Adults. JAMA Netw Open. 2024;7(10):e2441970–e2441970.

9.	Zhang B, Thacker D, Zhou T, Zhang D, Lei Y, Chen J, et al. Post-Acute Cardiovascular Outcomes of COVID-19 in Children and Adolescents: An EHR Cohort Study from the RECOVER Project. medRxiv. 2024;

10.	Lu Y, Tong J, Zhang D, Chen J, Li L, Lei Y, et al. Does SARS-CoV-2 Infection Increase the Frequency and Risk of Neuropsychiatric and Related Conditions? Findings from Difference-in-Differences Analyses.

11.	Li L, Zhou T, Lu Y, Chen J, Lei Y, Wu Q, et al. Post-acute and Chronic Kidney Function Outcomes of COVID-19 in Children and Adolescents: An EHR Cohort Study from the RECOVER Initiative. medRxiv. 2024;

12.	Zhang D, Stein R, Lu Y, Zhou T, Lei Y, Li L, et al. Pediatric Gastrointestinal Tract Outcomes During the Postacute Phase of COVID-19. JAMA Netw Open. 2025;8(2):e2458366–e2458366.

13.	Schuemie MJ, Hripcsak G, Ryan PB, Madigan D, Suchard MA. Empirical confidence interval calibration for population-level effect estimation studies in observational healthcare data. Proceedings of the National Academy of Sciences. 2018;115(11):2571–7.

14.	Schuemie MJ, Ryan PB, DuMouchel W, Suchard MA, Madigan D. Interpreting observational studies: why empirical calibration is needed to correct p‐values. Stat Med. 2014;33(2):209–18.
